# Regional excitability, not epileptic pathology, drives stimulation-evoked interictal spike increases

**DOI:** 10.64898/2026.05.21.26353811

**Authors:** Carlos A. Aguila, Zican Zhuo, Sarah B. Lavelle, William K.S. Ojemann, Juri Kim, Katherine G. Walsh, Sasan Sedighi Mournani, Alfredo Lucas, Nishant Sinha, Odile Feys, Brittany H. Scheid, Kathryn A. Davis, Brian Litt, Erin C. Conrad

**Affiliations:** Department of Bioengineering, School of Engineering & Applied Sciences, University of Pennsylvania, Philadelphia, PA 19104, USA; Center for Neuroengineering and Therapeutics, University of Pennsylvania, Philadelphia, PA 19104, USA; Department of Neurology, Perelman School of Medicine, University of Pennsylvania, Philadelphia, PA 19104, USA; Department of Biostatistics and Epidemiology, University of Pennsylvania, Philadelphia, PA 19104, USA; Cadence Neuroscience, Inc., Redmond, WA, USA

## Abstract

**Objective:** Interictal spikes have been proposed as a biomarker for both localizing seizure onset zones (SOZ) and tracking changes in seizure risk with neurostimulation in patients with drug-resistant epilepsy. Electrical stimulation can modulate spike rates acutely, and it has been proposed that measuring this modulation can help localize the SOZ. However, it is unclear whether stimulation-induced spike rate changes reflect epilepsy-specific pathology in the stimulated network or simply intrinsic regional excitability, which limits our understanding of their utility in epilepsy surgery planning.

**Methods:** We analyzed low-frequency stimulation (LFS; 1 Hz) applied during a clinical seizure-induction protocol systematically targeting multiple brain regions in 43 patients with drug-resistant epilepsy undergoing intracranial EEG monitoring. A validated, automated spike detector was used to quantify pre-, during-, and post-stimulation spike rates. We tested whether the stimulation-evoked spike rate response (i) tracks the expected change in seizure risk from a seizure induction protocol, (ii) varies with anatomical stimulation site and epilepsy localization, (iii) localizes the SOZ beyond baseline spike rate, and (iv) is accompanied by changes in spike morphology.

**Results:** Nearby LFS acutely increased spike rates in high-spiking channels (inter-stimulation median 2.25 vs. during-stimulation 4.25 spikes/min; p < 0.001), with effects attenuating with distance and resolving within approximately 30 seconds of stimulation offset. Mesial temporal lobe stimulation produced the largest increase in nearby spike rates relative to temporal neocortex and other cortex (Kruskal-Wallis p = 0.003), but this effect did not differ between patients with and without mesial temporal lobe epilepsy. A random forest classifier incorporating stimulation-evoked modulation features achieved an AUC of 0.787, comparable to a resting-state spike model (AUC 0.747; DeLong p = 0.81), indicating that stimulation-evoked spike changes do not add localizing information beyond resting-state spike rates. Stimulation produced a small but significant shift in spike morphology toward broader, higher-amplitude discharges (PERMANOVA p < 0.001), consistent with recruitment of a broader neuronal population.

**Significance:** LFS-evoked increases in interictal spike rates reflect intrinsic regional excitability, greatest in the mesial temporal lobe, rather than epilepsy-specific pathology, and do not improve SOZ localization over resting-state spike rates. These results argue against using the change in spikes with stimulation to localize the SOZ. On the other hand, the transient spike rate increase induced by a pro-epileptic protocol supports the acute change in spike rate as a biomarker of stimulation’s effect on seizure risk, with potential to guide parameter selection for epilepsy neuromodulation.

**Key Points:**

- 1 Hz low-frequency stimulation performed for seizure induction acutely increases interictal spike rate in nearby cortex
- Mesial temporal structures show disproportionately large stimulation-evoked spike responses, independent of whether the patient has mesial temporal lobe epilepsy.
- Although stimulation-induced spike rates are higher in the seizure onset zone, induced spikes do not localize the seizure onset zone better than baseline spikes.
- Spikes during stimulation have different morphology than spikes at rest, supporting recruitment of a broader neuronal population.

## Introduction

Epilepsy is a common and often disabling neurological disorder characterized by spontaneous seizures. Although seizures define the clinical disease, the underlying process reflects a broader instability of neural networks^1–3^, in which small perturbations can shift circuits toward or away from seizures. Clinicians exploit this sensitivity using electrical stimulation to probe epileptic networks in the epilepsy monitoring unit (EMU) in patients undergoing intracranial EEG (iEEG) recording for epilepsy surgery planning. This procedure relies on the assumption that the local cortical response to stimulation reflects pathological excitability of the underlying epileptic network, revealing both epileptogenicity and the stimulation-induced change in seizure risk.

Stimulation can induce seizures^4^, alter connectivity and spectral activity^5–7^, and modulate interictal spikes — brief epileptiform discharges that occur between seizures. Among proposed stimulation-induced biomarkers, interictal spikes are particularly attractive. Spikes are a biomarker of epilepsy severity^8^, decline with successful chronic neuromodulation^9,10^, and occur much more frequently than seizures, making them well-suited to quickly probe epileptic networks. Two applications of stimulation-induced spikes have been proposed for surgical planning. First, changes in spike rate with stimulation may track changes in seizure risk — such that a reduction in interictal spikes could signal an effective chronic neuromodulation parameter set or target location^11–14^. Second, because spikes reflect epileptic hyperexcitability, stimulation-evoked increases in interictal spikes and other epileptiform activity may localize the seizure onset zone (SOZ)^4,15–17^.

Two gaps in the literature limit both applications: small sample sizes, often driven by reliance on visual spike detection, and a lack of systematic exploration of stimulation locations, which confounds our interpretation of stimulation responses. For example, the mesial temporal lobe (MTL) exhibits lower seizure thresholds than the rest of the cortex in both humans and animal models^18–20^, with 1 Hz stimulation of mesial temporal or insular contacts generating seizures in 47% of trials compared with 5% elsewhere^21^. This raises a crucial question: do stimulation-evoked spike increases reflect underlying pathology or normal regional hyperexcitability? Resolving this question requires comparing the stimulation response across regions and across epileptic localizations.

Here, we studied how low-frequency stimulation (LFS) affects interictal spike rates and how this varies by location. We leveraged a dataset of 1 Hz LFS applied during iEEG as part of a clinical seizure induction protocol that systematically stimulated several brain regions in each patient^22^. We tested two hypotheses. First, that spike rates increase with a seizure induction stimulation protocol, consistent with spikes tracking seizure risk. Second, that the magnitude of stimulation-evoked spike rate changes localizes the SOZ. With this question, we seek to address whether stimulation-induced spike rate changes reflect epilepsy-specific pathology or intrinsic regional excitability.

## Materials and Methods

### Patient Selection and Clinical Determinations

We retrospectively analyzed 62 sequential patients with drug-resistant epilepsy who underwent iEEG monitoring and low-frequency electrical stimulation at the Hospital of the University of Pennsylvania (HUP) between 2021 and 2025. SOZ localization was determined by the clinical team in a surgical conference following review of all iEEG data. Given evidence that the mesial temporal lobe structures are hyperexcitable^18^, we classified SOZ localizations into mesial temporal lobe epilepsy (MTLE) vs other. The HUP IRB (protocol # 821778) approved the research and all patients provided written informed consent to retrospective analysis of their data.

### Intracranial Recording

Patients were implanted with subdural grid, strip, and/or depth electrodes (Ad-Tech Medical Instrument Corporation, Oak Creek, WI) based on clinical needs. Signals were recorded using a Natus Quantum LTM Amplifier (Natus Medical Incorporated, Middleton, WI) at a sampling frequency ranging from 512-2048 Hz. Signals were referenced to an electrode distant from the presumed epileptogenic tissue. Electrode localization was performed for all patients according to our published protocol^23^.

### Stimulation Protocol

LFS was delivered as part of a clinical protocol to induce seizure using 1 Hz stimulation frequency, 300–500 μs pulse width, 3 mA current (11.1–18.5 μC/cm²), and 30 s train duration (**Fig. 1A**). This protocol has a rate of seizure induction of 39.5% of patients at our center, similar to previous reports of stimulation-induced seizures^4,21,24^. Clinicians waited a median (IQR) of 17.34 s (13.82 - 23.38 s) between delivering trains of stimuli. Adjacent bipolar pairs in neural tissue were stimulated unless they demonstrated high-amplitude artifact or if time constraints from clinical care necessitated sampling every other pair. Stimulation sessions typically were halted if a clinical seizure with impaired consciousness was induced. Stimulation sessions lasted a median of 83.3 (IQR: 70.2 – 100.0) minutes and were performed at a median of 16.2 (IQR: 3.2 – 24.7) hours after the onset of electrode recording, while patients were on near-home doses of anti-seizure medications.

**Figure 1.**
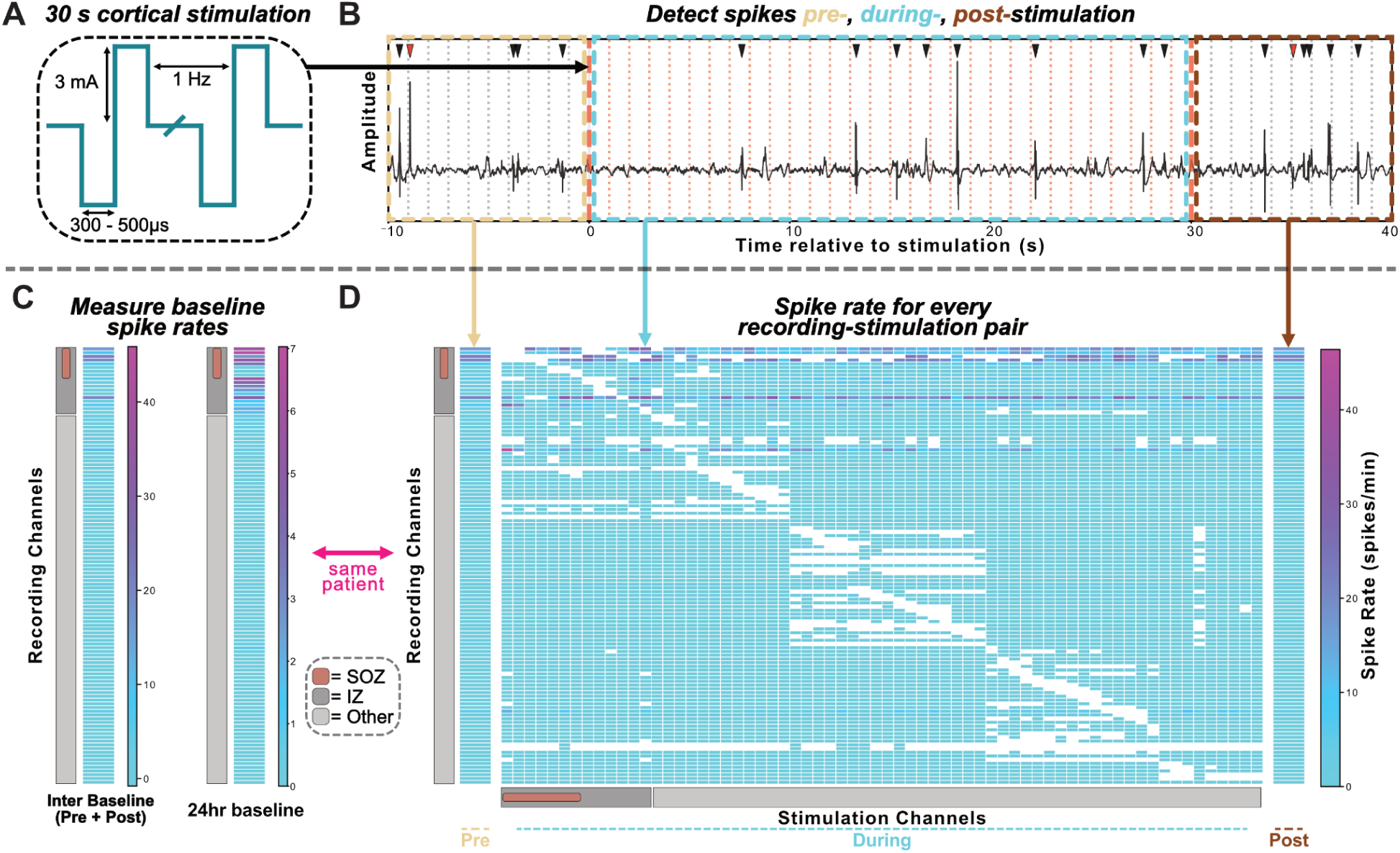
Analysis pipeline for quantifying stimulation-evoked changes in interictal spike rate. (A) Schematic of the low-frequency stimulation (LFS) protocol: biphasic pulses delivered at 1 Hz, 3 mA, with pulse widths of 300–500 μs for 30 seconds. (B) Representative iEEG trace from a single recording channel illustrating the three analysis epochs: pre-stimulation (yellow), during-stimulation (teal), and post-stimulation (brown). Black arrowheads indicate spikes included in the rate calculation; red arrowheads indicate spike detections that fell within the 95 ms post-pulse exclusion window and were rejected to avoid misclassifying cortico-cortical evoked potential (CCEP) components as interictal spikes. Vertical dotted lines mark individual stimulation pulses or sham stimulations. (C) Baseline spike rate heatmaps for all recording channels in a representative patient. Inter-stimulation baseline (left) is computed from the combined pre- and post-stimulation epochs across all trials for a single recording electrode; 24-hour baseline (right) is derived from spike detection in ±12 hr windows to the stimulation session. Channels are color-coded by clinical classification: seizure onset zone (SOZ; blue), irritative zone (IZ; orange), and non-epileptiform channels (gray). (D) Spike rates for every recording–stimulation channel pair across all patients, organized by pre-stimulation, during-stimulation, and post-stimulation epochs. Each row is a recording channel; columns correspond to stimulation channels from the same patient (indicated by the pink bracket). Color encodes spike rate (spikes/min); white cells indicate pairs excluded because of heavy artifact.

### 24-hour baseline spike rate distribution

To quantify baseline interictal spiking independent of stimulation, we measured the spike rate within +/- 12 hours of the stimulation session using a previously validated, in-house spike detector designed for EMU recordings.^25–27^ Within each window, we detected spikes in 1,000 randomly sampled non-overlapping 30-second clips. We defined the 24-hour baseline spike rate as the mean spike rate across all sampled clips, yielding a stable channel-level baseline distribution used in downstream analyses **(Fig. 1C)**.

### Spike detection parameter tuning and validation

Given that stimulation may affect the accuracy of automated spike detection, we performed additional validation to ensure that the spike detector was not biased in sensitivity or false positive rates comparing the inter-stimulation versus during-stimulation periods. An epileptologist (E.C.C.), blinded to automated detections, manually labeled spikes in 500 independent 50s clips (10s pre-, 30s during-, 10s post-stimulation) sampled across stimulation/recording channel pairs with spike activity detected from an unoptimized detector. These 500 clips were randomly divided into a training set (350 clips) and a held-out test set (150 clips). Spike detector hyperparameters were selected from the training set to maximize sensitivity. The resulting fixed parameter set was then validated on the held-out test set to confirm that detector performance was not biased across pre-, during-, and post-stimulation windows. This final parameter set was applied to all subsequent analyses. To exclude patients with low spike detector accuracy in the peri-stimulation data, E.C.C. further visually reviewed 20 random detections per patient; those with a positive predictive value <70% were excluded from subsequent analyses.

### Peri-stimulation spike detection

To quantify stimulation-induced changes in spiking, we compared spike rates measured during stimulation against those measured between stimulation trains.

#### Division into pre-, during-, and post-stimulation windows

Given that clinicians usually waited at least 10 seconds from finishing one stimulation train to starting the next, we analyzed each 1-Hz stimulation train as a single 50 s epoch comprising 10 s pre-stimulation, 30 s during stimulation, and 10 s post-stimulation **(Fig. 1B)**. Stimulation trains that elicited a seizure were excluded from analysis, removing the entire 50 s epoch (pre-, during-, and post-stimulation windows) for that trial. Trials delivered after a stimulation-induced seizure in the same session were retained when stimulation was continued; this applied to 2 patients (21 and 5 post-seizure trials, respectively). Because stimulation produces a high amplitude artifact that may be falsely detected as a spike, we removed stimulation artifacts by linearly interpolating across a 0.06 s window surrounding the stimulation event^28^. To ensure this interpolation process did not bias spike detection to be higher or lower during the stimulation window periods, we applied the same interpolation procedure in the pre- and post-stimulation windows at 1 s spaced “sham” timestamps, matched to the timing structure of the true stimulation train. The spike detector was run on the full 50 s clip, and detections were subsequently assigned to the during- and inter-stimulation periods for rate calculations.

#### Spike rate calculations

Both during-stimulation and inter-stimulation spike rates were computed using the same procedure. After artifact interpolation, each 1 s cycle contributed a 0.94 s analyzable segment. To avoid falsely detecting stimulation-evoked responses (e.g., CCEPs) as spikes, we excluded detections occurring within 95 ms of each stimulation pulse (or sham, in the case of the pre- and post- periods)^29,30^. Spike rate was computed as the number of remaining detections divided by the total non-excluded duration in the corresponding window (**Fig. 1D**).

“During-stimulation” spike rates were defined as those occurring during the 30 s stimulation train. Inter-stimulation baseline was estimated by pooling spike detections from the 10 s pre-stimulation and 10 s post-stimulation windows (**Fig. 1C**). To prevent double-counting spikes across consecutive trials, we identified any overlap between a trial’s post-stimulation window and the subsequent trial’s pre-stimulation window; spikes falling within the overlapping region were counted only once before pooling.

### Selection of recording and stimulation channels

To study the effect of stimulation on the epileptic network, we primarily analyzed the effect of stimulation on spikes in the irritative zone (IZ), defined as channels whose 24-hour baseline spike rate was above 1.0 spikes/min (87th percentile across the full cohort). We chose these channels rather than the clinician-defined SOZ for our primary analysis because the SOZ often consists of a very small number of channels, too few for robust estimates of spike rates, and the irritative zone is believed to be functionally connected to the epileptic network^31,32^. IZ and SOZ designations overlapped in all but 3 patients for whom both sets were defined. We also performed a secondary analysis testing the stability of our primary results when analyzing the SOZ as opposed to the IZ.

To isolate local stimulation effects, we primarily analyzed stimulations occurring within 40 mm of the recording site, which we refer to as "nearby" stimulation, consistent with prior work^11^.

### Machine Learning to predict channels in the SOZ

Prior work has demonstrated that single-pulse electrical stimulation can elicit *delayed responses* — late (> 100ms after stimulation) epileptiform discharges evoked by cortical stimulation — that preferentially arise when stimulating within or near the SOZ, suggesting that stimulation-evoked epileptiform activity carries localizing information^16,17^. Because delayed responses morphologically resemble spontaneous interictal spikes and likely share their generators^33,34^, we used stimulation-induced spike rates as a proxy. We therefore tested whether the modulation in spike rate with stimulation improves SOZ localization beyond what is captured by the 24-hour baseline spike rate alone. For each channel, features included (i) the 24-hour baseline spike rate and (ii) two directional stimulation-response metrics: outward modulation, defined as the change in spike rate observed in nearby recording channels (< 40 mm) when stimulating the target channel, and inward modulation, defined as the change in spike rate observed in the target channel when stimulating nearby channels. Together, these features capture both how a candidate SOZ channel perturbs its local network when stimulated and how strongly it is modulated by stimulation delivered to adjacent tissue.

Classification was performed using a random forest model predicting whether each channel was a SOZ with 24-hour baseline spike rate, outward modulation, and inward modulation as features, and was trained, tested, and tuned using a nested, two-loop leave-one-patient-out cross-validation framework to avoid bias and information leakage (**Supplemental Materials**).

### Comparison of spike morphology

We hypothesized that stimulation may affect *which* neurons produce interictal spikes, rather than just the frequency of spiking of the same underlying neurons. To test this hypothesis, we compared the spike morphology during and between stimulation, given that spike morphology reflects the underlying generating ensemble^25,35–38^. We computed eight morphological features for each detected spike: sharpness, width, line length, slow wave width, slow wave amplitude, rise slope, decay slope, and average amplitude (detailed computation is in **Supplemental Materials**; spike morphology calculations were performed using our previously validated method)^25,39,40^. Pairwise Euclidean distances between spike features were converted to a Gower-centered Gram matrix via double-centering^41,42^, which reformulates pairwise distances as inner products and enables the sums-of-squares decomposition required by PERMANOVA. To determine whether stimulation conditions (during vs. inter-stimulation) explained a significant portion of multivariate morphological variance after accounting for known sources of confounding, we applied a stratified permutation MANOVA (PERMANOVA). Variance was partitioned sequentially (Type I SS), entering terms in the following order: patient identity, stimulation channel, recording channel, and finally condition. This ordering ensures that between-patient differences and electrode-level effects are accounted for before testing the effect of condition, preventing spurious significance driven by these nuisance factors.

### Statistical analyses

To compare two independent groups, we report Mann–Whitney U tests (α = 0.05) and Cliff’s delta (d) as a measure of effect size. Paired differences were measured using the Wilcoxon signed-rank test, and multi-group comparisons were evaluated with Kruskal–Wallis tests. For the PERMANOVA, statistical inference was performed using a stratified permutation test with 999 permutations (**Supplemental Materials**). Model performance was characterized using receiver operating characteristic (ROC) curves with corresponding area under the curve (AUC), as well as precision–recall curves, balanced accuracy, and average precision (AP). Statistical comparisons between AUCs were performed using the DeLong method. All analyses were performed in Python 3.12.0. For all statistical assessments, the significance threshold was set at α = .05, with Bonferroni correction applied for multiple comparisons where appropriate. In all plots, p-values are denoted using the following abbreviations: ns – p >= 0.05 (not significant), * – p < 0.05, ** – p < 0.01, *** – p < 0.001, **** – p < 0.0001.

## Results

### Patient Information

Of 62 patients, 15 were excluded due to a spike detector PPV < 0.7 from the peri-stimulation dataset. Excluded patients had lower detected spike rates than included patients, likely explaining the lower PPV, but were otherwise similar in clinical characteristics (**Supplemental Table 1; Supplemental Fig. S3-S4**). An additional 4 patients were excluded for lacking any detected spikes in either the inter- or during-stimulation windows. The remaining 43 patients are summarized in Table 1.

**Table 1.**
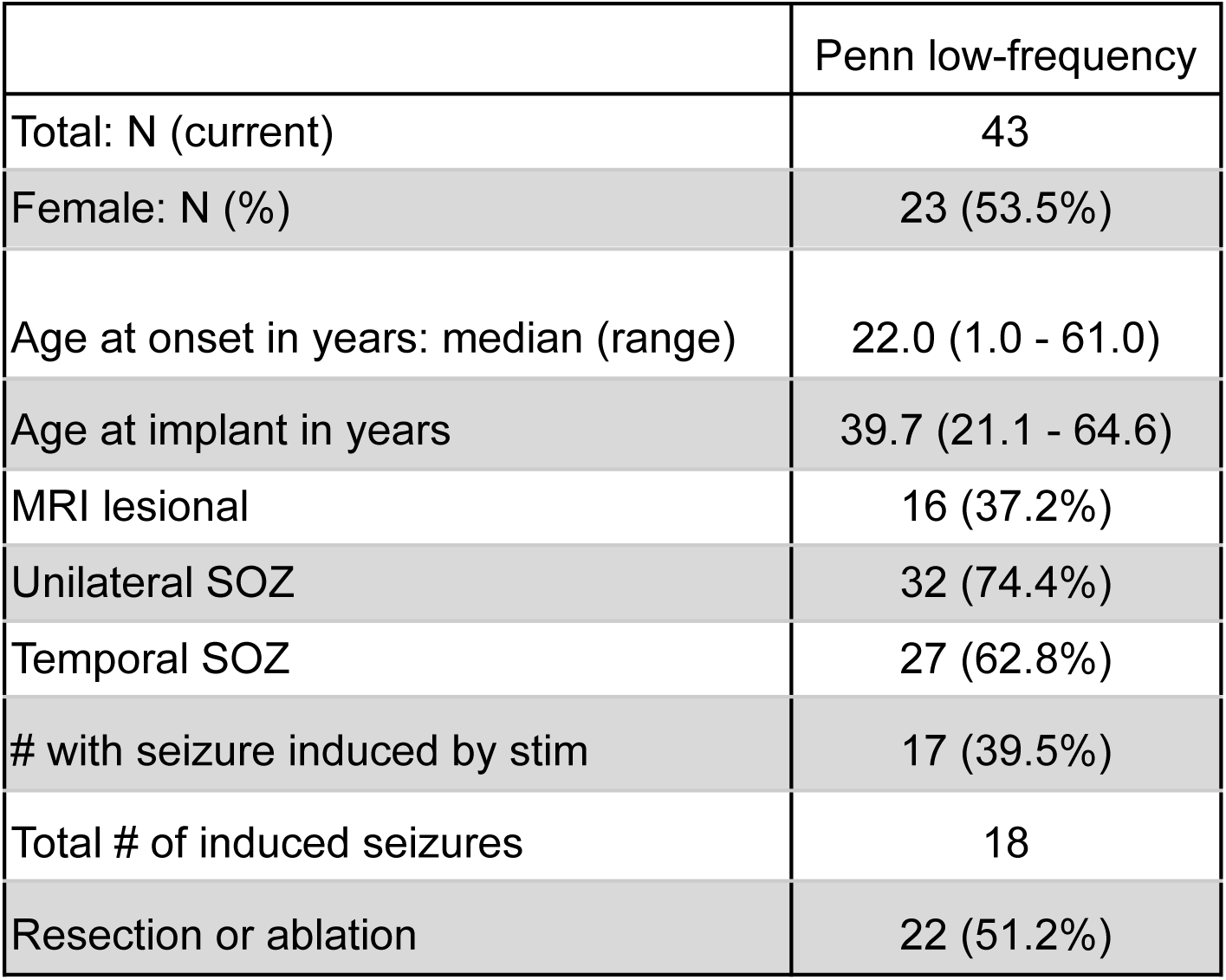
Patient demographics (n = 43). Abbreviations: SOZ = seizure onset zone.

|  | Penn low-frequency |
| --- | --- |
| Total: N (current) | 43 |
| Female: N (%) | 23 (53.5%) |
| Age at onset in years: median (range) | 22.0 (1.0 - 61.0) |
| Age at implant in years | 39.7 (21.1 - 64.6) |
| MRI lesional | 16 (37.2%) |
| Unilateral SOZ | 32 (74.4%) |
| Temporal SOZ | 27 (62.8%) |
| # with seizure induced by stim | 17 (39.5%) |
| Total # of induced seizures | 18 |
| Resection or ablation | 22 (51.2%) |

Spike detector sensitivity and precision were similar across the pre-, during-, and post-stimulation phases, suggesting that stimulation artifact did not bias automated detection and corresponding spike rate estimates (**Supplemental Fig. S5**). The correlation in spike rate across channels between the 24-hour baseline and inter-stimulation period was a median (IQR) of 0.57 (0.38 - 0.73) across patients, suggesting that spike rates in between stimulation trains are similar to those during the baseline period (**Supplemental Fig. S6**).

### Nearby stimulation increases spike rates in high-spiking channels

Out of 43 patients, 35 had electrode contacts in the IZ (24-hour baseline spike rate > 1.0 spikes/min). The effect of stimulation on spike rates diminished with distance between the stimulating and recording contact (**Fig. 2A-B**).

**Figure 2.**
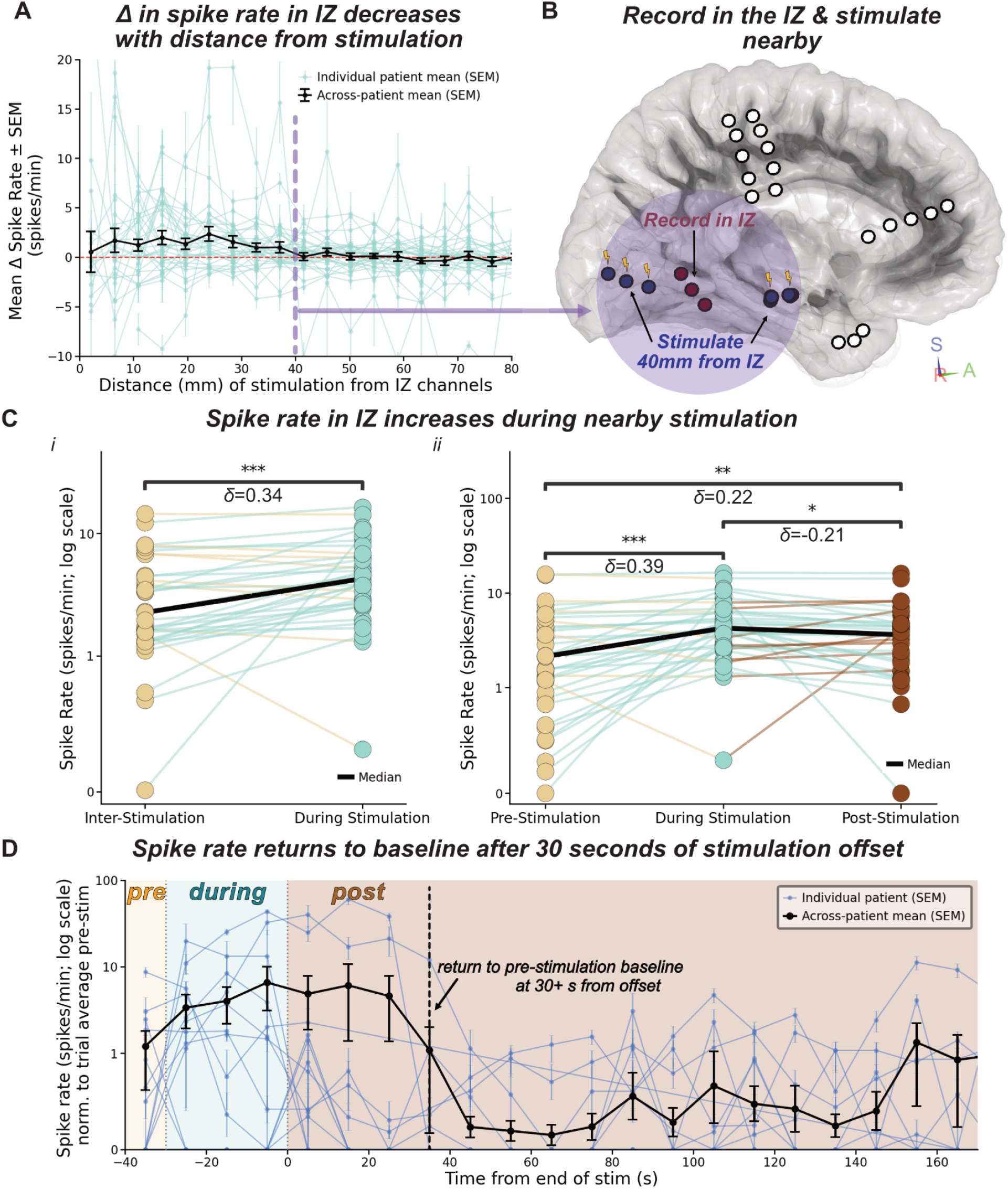
Low-frequency stimulation increases spike rate in the irritative zone in a distance-dependent manner. (A) Mean change in IZ spike rate (spikes/min ± SEM) as a function of stimulation-to-recording channel distance (n = 35 patients with IZ electrodes). Teal lines represent individual patient means; black line with error bars represents the across-patient mean. The dashed purple vertical line at 40 mm marks the boundary used to define "nearby" stimulation. The stimulation-evoked increase in spike rate decays monotonically with distance, approaching zero beyond 40 mm. (B) Schematic illustrating the recording and stimulation configuration. Red contacts indicate IZ recording channels; blue contacts indicate stimulation delivered within 40 mm of the IZ. (C) IZ spike rate (spikes/min, log scale) during inter-stimulation and during-stimulation periods, shown for all nearby stimulation contacts (n = 35 patients with IZ electrodes; i) and stratified by pre-, during-, and post-stimulation windows (n = 35; ii). Each dot represents one patient; black lines indicate the cohort median. Colored lines indicate patient tracked spike rate and the color denotes which window had a larger spike rate. (D) Time course of IZ spike rate recovery following the last stimulation trial, restricted to patients without stimulation-induced seizures (n = 15). Panel (i) shows IZ spike rate normalized to the trial average pre-stimulation baseline as a continuous function of time from the last stimulation pulse. Shaded regions indicate the pre-stimulation (yellow), during-stimulation (teal), and post-stimulation (brown) epochs. Blue lines represent individual patients (± SEM); black line represents the across-patient mean (± SEM). The dashed purple vertical line marks the time at which the across-patient mean visually appears to return to pre-stimulation baseline (∼30 s post-offset).

Spike rate in the IZ was higher during nearby stimulation (median 4.25; IQR 2.71 – 6.40 spikes/min) than during the inter-stimulation baseline (2.25; 1.42 – 4.48 spikes/min) (Wilcoxon signed-rank test, p < 0.001; Cliff’s δ = 0.344; **Fig. 2C-i**). Stratified by stimulation window, spike rate increased significantly from pre-stimulation (2.14; 0.93–4.73 spikes/min) to during-stimulation (4.25; 2.71–6.40 spikes/min; p_corrected_ < 0.001, δ = 0.39). The post-stimulation window (3.6; 1.67–4.82 spikes/min) remained elevated above the pre-stimulation baseline (p_corrected_ < 0.01, δ = 0.22), but was significantly lower than the during-stimulation peak (p_corrected_ < 0.05, δ = −0.21; **Fig. 2C-ii**). These results support that 1 Hz LFS with our parameters produces an acute, spatially constrained increase in interictal spike rate in nearby high-spiking tissue. A secondary analysis restricted to SOZ channels showed a similar inter-to-during and pre-to-during increases in spike rate (**Supplemental Fig. S7**).

To characterize the time course of spike rate recovery, we conducted a descriptive analysis of IZ spike rate normalized to the pre-stimulation baseline, measured from the time of the last stimulation pulse during each patient’s stimulation session, restricted to patients without stimulation-induced seizures to avoid confounding the recovery trajectory with post-ictal suppression (n = 15; **Fig. 2D**). Across patients, the mean spike rate was elevated during the stimulation train and declined progressively after offset, with visually apparent return to baseline within ∼ 30 seconds of stimulation offset. Our modest sample size precluded inferential testing of the bin-by-bin recovery trajectory. A limited paired comparison restricted to the post-stimulation window (**Supplemental Fig. S8**) was consistent with return to pre-stimulation baseline by 60 seconds after stimulation offset. These observations are consistent with a transient stimulation-evoked increase in excitability that self-resolves within one minute.

### Stimulating in the MTL is associated with the greatest increase in spikes

We next asked whether the effect on excitability depends on the anatomical location of stimulation. We categorized the stimulation region as mesial temporal lobe (MTL), temporal neocortex (TN), or other cortex (OC), and measured spike rate changes in recording channels within 40 mm of the stimulation contact, as illustrated schematically in **Fig. 3A**. Comparing inter-stimulation to during-stimulation spike rates within each region, only MTL stimulation produced a significant increase in median spike rate (MTL: 0.89 to 1.35 spikes/min, p_corrected_ < 0.001; TN: 0.78 to 0.59, ns; OC: 0.53 to 0.51 spikes/min, ns; **Fig. 3B-i**). The change in spike rate varied across stimulation regions (MTL: median 0.33; IQR −0.02 – 0.83 spikes/min, n = 41; TN: −0.10; −0.37 – 0.22 spikes/min, n = 42; OC: −0.01; −0.22 – 0.29 spikes/min, n = 43; Kruskal–Wallis, p = 0.003; **Fig. 3B-ii**). Post hoc comparison revealed that MTL stimulation produced significantly larger increases in nearby spike rate than TN stimulation (p_corrected_ = 0.003, δ = 0.42). These results indicate that the excitatory response to 1 Hz stimulation is anatomically specific, with nearby tissue responding most strongly when the stimulation is performed in the mesial temporal lobe.

**Figure 3.**
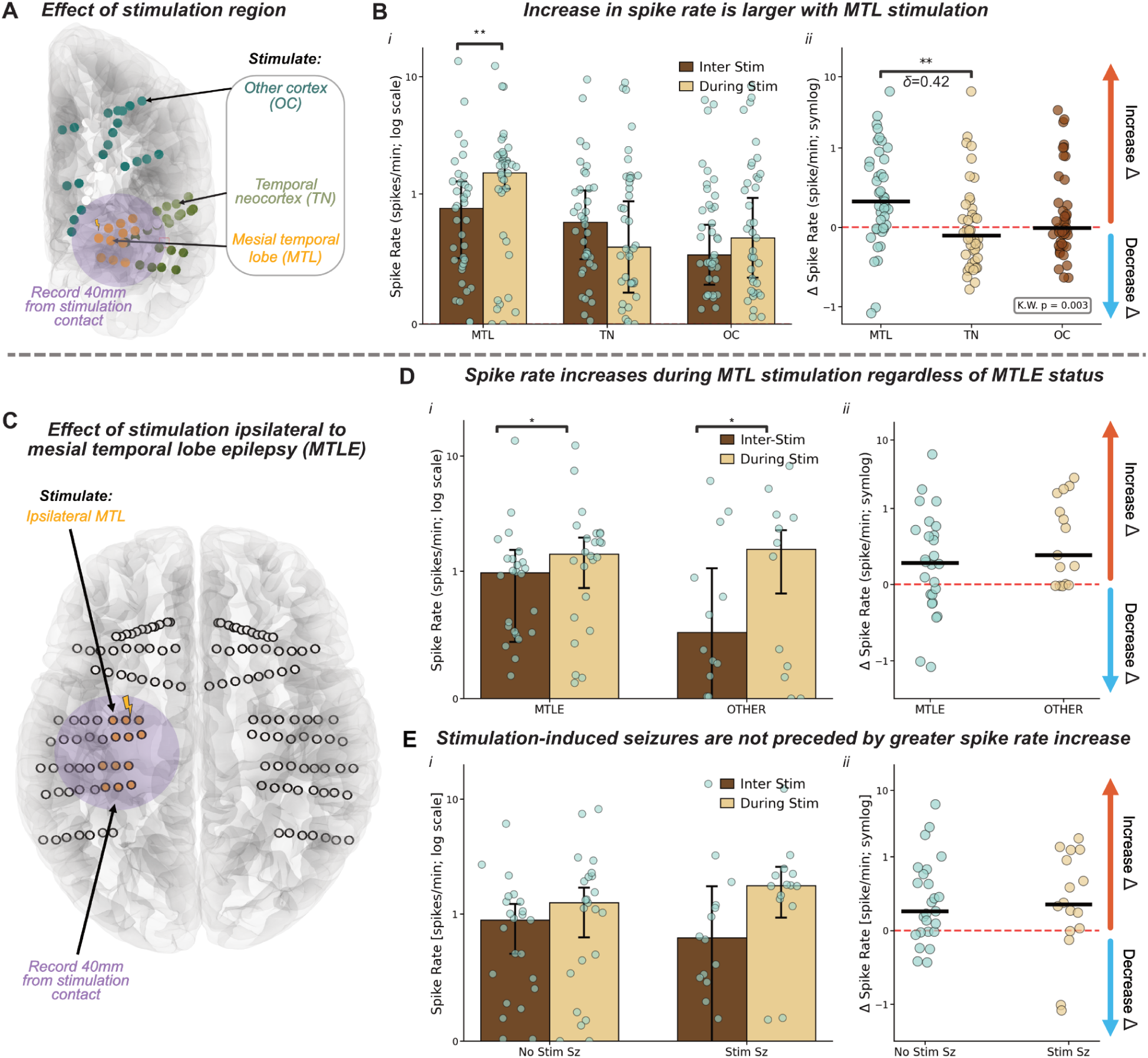
MTL stimulation produces the largest increase in nearby spike rate, independent of epilepsy localization and seizure induction. (A) Brain schematic illustrating the three stimulation region categories: mesial temporal lobe (MTL, orange), temporal neocortex (TN, olive), and other cortex (OC, teal). Spike rates are recorded from contacts within 40 mm of the stimulation site. (B) Spike rate (spikes/min, log scale) during inter-stimulation and during-stimulation periods stratified by stimulation region (MTL: n = 41 patients with MTL stimulation TN: n = 42; OC: n = 43; i), and the corresponding distribution of change in spike rate per patient on a symmetric log (symlog) scale, which displays both positive and negative values by transitioning from linear near zero to logarithmic at larger magnitudes (ii). Each dot represents one patient; bars indicate the median. MTL stimulation produced significantly larger increases in nearby spike rate than TN or OC stimulation (K-W p = 0.006; Cliff’s δ = 0.42). (C) Brain schematic (axial view) illustrating the ipsilateral MTL stimulation configuration in patients with mesial temporal lobe epilepsy (MTLE), with recordings from contacts within 40 mm of the stimulation site. (D) Spike rate during inter-stimulation and during-stimulation periods for MTLE patients versus those with other epilepsy localizations (MTLE: 1.00 → 1.41 spikes/min, p_corrected_ = 0.04, n = 26; non-MTLE: 0.50 → 1.34 spikes/min, p_corrected_ = 0.007, n = 15), and the corresponding Δ in spike rate distributions on a symlog scale (ii). Spike rate increased significantly during MTL stimulation in MTLE patients (p < 0.05) but not in non-MTLE patients, though the Δ spike rate did not differ significantly between groups. (E) Spike rate during inter-stimulation and during-stimulation periods in patients with versus without stimulation-induced seizures (seizure: n = 16, no seizure: n = 25), and the corresponding change in spike rate distributions on a symlog scale (ii). No significant differences were observed between groups, indicating that stimulation-induced seizures are not preceded by a greater increase in spike rates.

### The effect of MTL stimulation on spike rate is independent of epilepsy localization

We next asked whether the excitatory response to MTL stimulation is exacerbated in patients with mesial temporal lobe epilepsy (MTLE). We analyzed MTL stimulation, averaging stimulations across both mesial temporal lobes in patients without MTLE and in patients with bilateral MTLE. We included only MTL stimulations ipsilateral to the SOZ in those with unilateral MTLE. We compared the corresponding change in spike rate between those with and without MTLE (**Fig. 3C**). As expected, MTLE patients had higher inter-stimulation spike rates near the MTL at baseline than non-MTLE patients (MTLE: median 0.99, IQR 0.53 – 1.28 spikes/min, n = 26; non-MTLE: 0.50, 0.28 – 1.19 spikes/min, n = 15), consistent with the MTL being near their seizure onset zone (**Fig. 3D-i**). However, the *increase* in spike rate from inter- to during-stimulation did not differ between epilepsy localizations (MTLE: 0.28, −0.14 – 0.70 spikes/min; non-MTLE: 0.39, −0.01 – 1.32 spikes/min; Mann-Whitney p = 0.17; **Fig. 3D-ii**). These results indicate that the acute excitatory response to MTL stimulation is not specific to patients with MTLE, and may reflect a general property of mesial temporal excitability rather than a pathological feature of the epileptic network.

We also compared the change in spike rate between the 16 patients who experienced seizures elicited by low-frequency MTL stimulation and 25 patients who did not have a seizure, excluding the stimulation periods that provoked the seizure. At contacts near the stimulation site, the inter-to-during spike rate change did not differ between patients with and without stimulation-induced seizures (**Fig. 3E**). This suggests that the excitatory change with stimulation may be similar regardless of whether stimulation induces a seizure.

### Stimulation-evoked spike rate changes do not improve SOZ localization beyond baseline spike rate

Prior work has shown that single-pulse electrical stimulation can elicit delayed responses that localize the SOZ**^15–17^**. We tested whether stimulation-evoked modulation of spike rate, captured as both outward modulation (change in spike rate in nearby channels when stimulating a candidate SOZ channel) and inward modulation (change in spike rate at a candidate channel when stimulating nearby; **Fig. 4A**), added predictive value for SOZ localization beyond the 24-hour baseline spike rate alone. A random forest classifier incorporating all three features achieved an AUC of 0.787 (95% CI: 0.737–0.832), compared to an AUC of 0.747 (95% CI: 0.685–0.800) for a baseline-only model (DeLong’s p = 0.81; **Fig. 4B**). Feature importance analysis confirmed that the 24-hour baseline spike rate dominated classification (importance = 0.564), with the two stimulation-response features contributing marginally (**Fig. 4C**). These results indicate that baseline interictal spiking captures the majority of localizing information and that stimulation-evoked spike modulation does not meaningfully improve SOZ prediction — consistent with the earlier finding that the stimulation-evoked spike rate increase does not vary with epilepsy localization.

**Figure 4.**
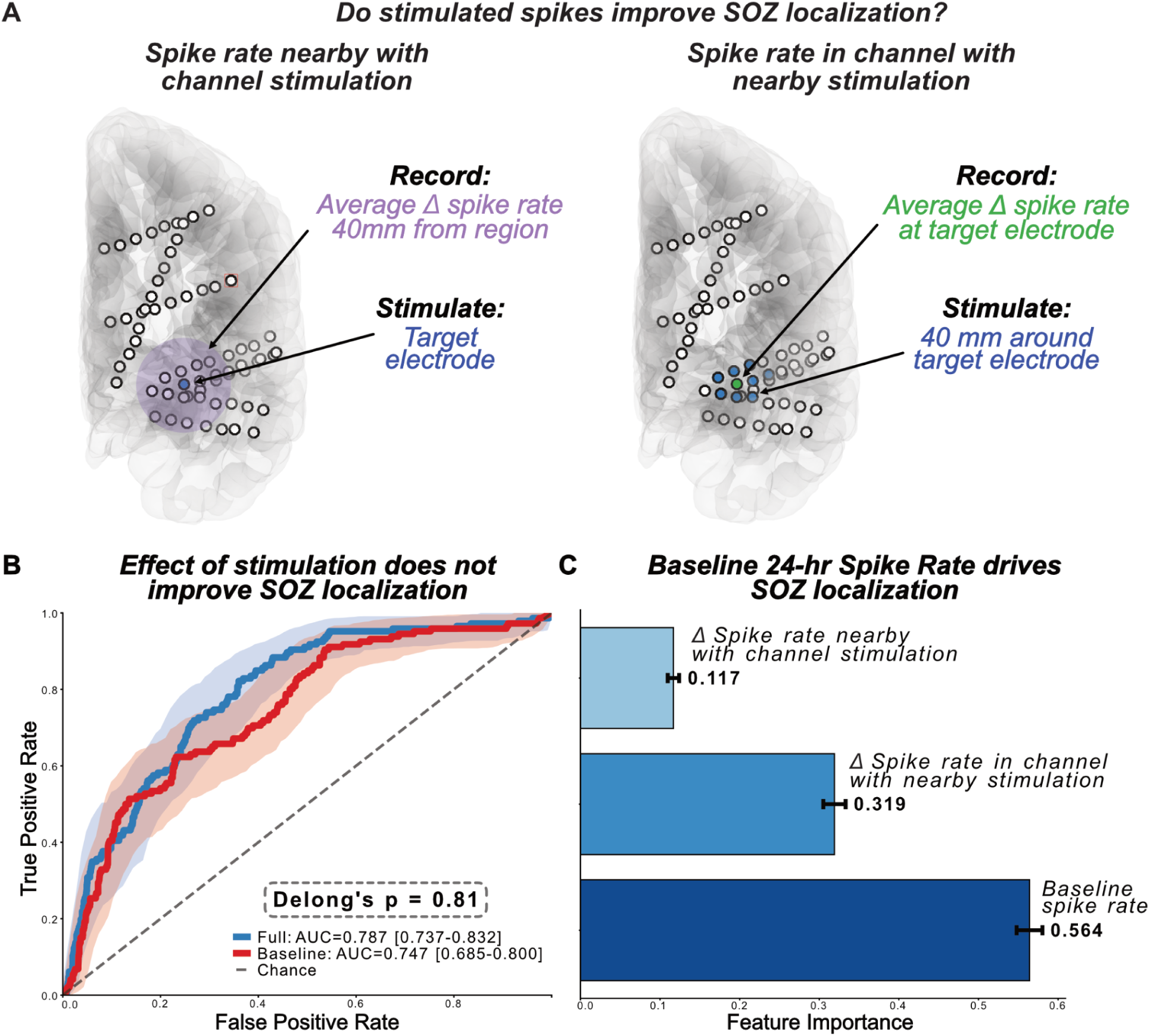
Stimulation-evoked spike rate changes do not improve SOZ localization beyond baseline spike rate alone. (A) Schematic of the two stimulation-based features used as inputs to the SOZ localization classifier. Left: the average Δ in spike rate in channels within 40 mm of a target electrode when that electrode is stimulated ("spike rate nearby with channel stimulation"). Right: the average Δ spike rate at a target electrode when nearby channels (within 40 mm) are stimulated ("spike rate in channel with nearby stimulation"). (B) ROC curves for the full classifier model (blue; AUC = 0.787 [0.737–0.832]) and the baseline-only model (red; AUC = 0.747 [0.685–0.800]). Shaded regions indicate 95% confidence intervals. The two models did not differ significantly in AUC (DeLong’s p = 0.81), indicating that including stimulation-evoked features did not meaningfully improve SOZ localization. (C) Feature importance estimates from the full classifier for each of the three input features. Baseline 24-hour spike rate was by far the strongest predictor (importance = 0.564), followed by Δ spike rate in channel with nearby stimulation (0.319) and Δ spike rate nearby with channel stimulation (0.117).

### Spike morphology is altered during stimulation

We next asked whether stimulation alters individual spike morphology rather than just spike rates, which would suggest recruitment of a broader neuronal population (**Fig. 5A**). A UMAP projection of residualized spike features, computed after removing patient and electrode contributions, shows during- and inter-stimulation spikes largely intermingled, with no clear visual separation between conditions, consistent with the small proportion of variance explained by the stimulation effect (**Fig. 5B**). Despite the small magnitude of the effect, the observed pseudo-F statistic (F = 56.9) far exceeded the null distribution generated by 999 stratified label permutations (p < 0.001; **Fig. 5C**), confirming a significant effect of stimulation condition on spike morphology. The stimulation effect accounted for 0.36% of total morphological variance, compared to 7.2% for patient identity, 4.2% for stimulating electrode, and 21.3% for recording electrode (**Fig. 5D**). Examining individual features, spikes occurring during stimulation were significantly broader (greater total width and slow wave width), with larger slow wave amplitude, line length, and peak amplitude, while sharpness was significantly reduced compared to inter-stimulation spikes (**Fig. 5E**). These morphological shifts are consistent with stimulation recruiting a broader population of neurons into individual interictal spikes, although this was a much smaller effect than the effect of recording location.

**Figure 5.**
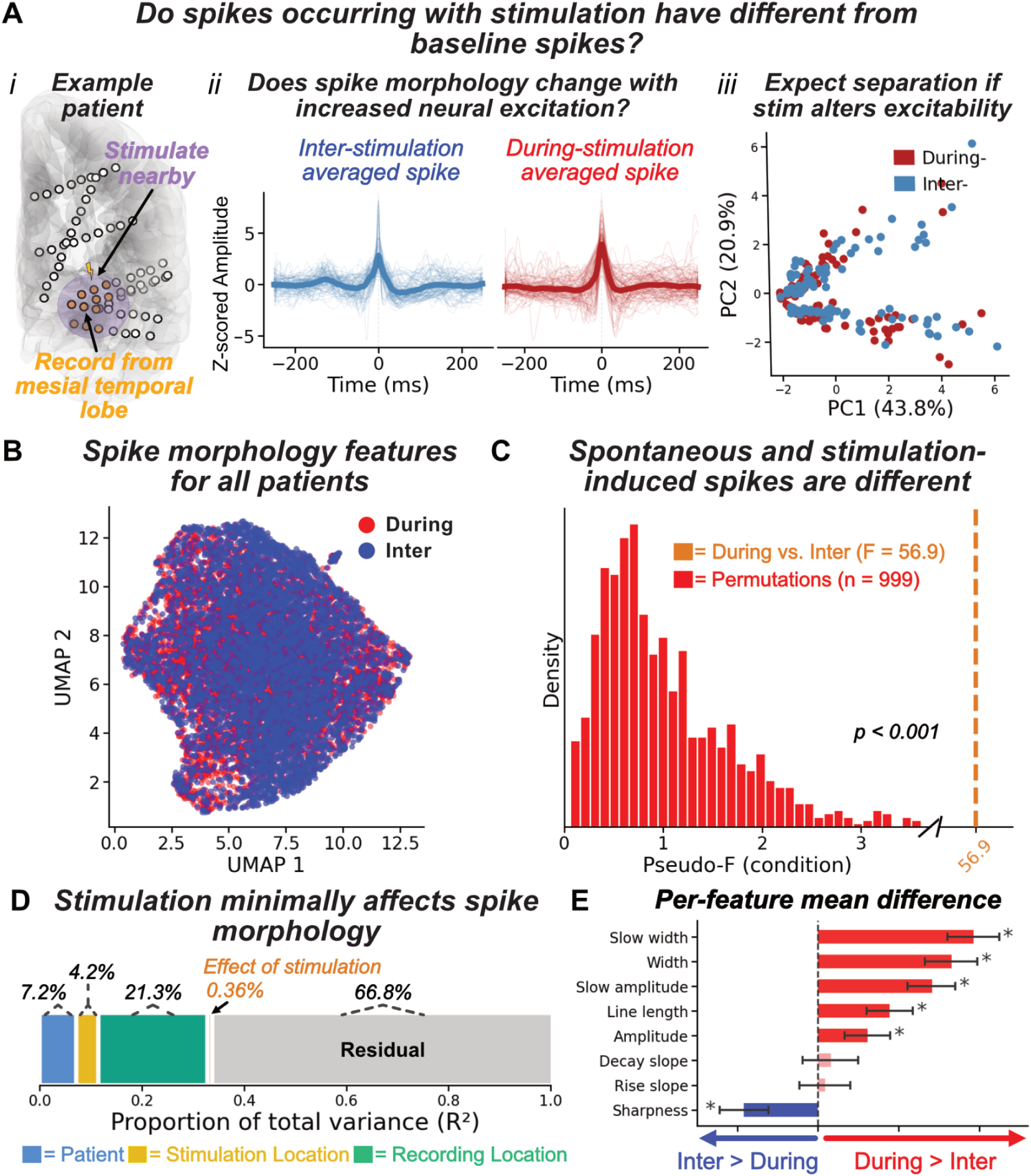
Stimulation transiently alters spike morphology, accounting for a small but significant proportion of variance. (A) Overview of the spike morphology analysis, illustrated for a single stimulation–recording electrode pair from a representative patient. Panel (i) shows the electrode configuration for this patient. Panel (ii) displays overlaid spike waveforms (z-scored amplitude) from inter-stimulation (blue) and during-stimulation (red) periods, illustrating an apparent change in waveform morphology. Panel (iii) depicts the expected feature-space separation between this patient’s inter- and during-stimulation spikes if stimulation alters excitability. (B) UMAP projection of residualized spike morphology features across all patients, color-coded by condition (blue = inter-stimulation, red = during-stimulation). The absence of clear cluster separation in the full UMAP reflects the dominance of patient- and electrode-level variance. (C) Null distribution of pseudo-F statistics from 999 permutations (red bars) compared to the observed pseudo-F for the stimulation condition (orange dashed line, F = 56.9), demonstrating a significant effect of the during vs. inter-stimulation condition on spike morphology (p < 0.001). (D) Proportion of total spike morphology variance attributable to patient (blue), electrode (green), stimulation condition (orange), and residual (gray) effects. The during vs. inter-stimulation effect accounts for 0.36% of total variance, compared to 7.2% for patients and 21.3% for recording electrodes. (E) Per-feature mean difference in spike morphology between during- and inter-stimulation periods (during minus inter), with error bars indicating SEM. Features marked with an asterisk differ significantly from zero after Bonferroni correction. During-stimulation spikes exhibit significantly greater slow wave width, total width, slow wave amplitude, and line length, alongside reduced sharpness.

## Discussion

We studied the effect of low-frequency stimulation on interictal spikes. Stimulation delivered as part of a seizure-induction protocol acutely increased spike rate in nearby high-spiking channels, with the effect decaying with distance. Mesial temporal lobe stimulation produced the largest increase in spikes, regardless of epilepsy localization, supporting that the increase is driven more by regional rather than epileptic excitability. The increase in spike rate with stimulation did not localize the seizure onset zone beyond baseline spike rate. Stimulation produced a small but significant shift in spike morphology, suggesting engagement of distinct or more broadly recruited spike generators. These findings suggest that the effect of stimulation on spike rates does not help localize seizure onset. On the other hand, the overall increase with a seizure induction stimulation protocol suggests that stimulation-induced spikes reflect the stimulation-induced change in seizure risk, supporting the use of stimulation-induced spikes as a biomarker of epilepsy neuromodulation.

### Low-frequency stimulation acutely increases spike rate, supporting that stimulation-induced changes in spikes reflect changes in seizure risk

We found that 1 Hz LFS in a seizure induction protocol acutely increased interictal spike rates in nearby high-spiking channels and at the seizure onset zone (SOZ). The mechanism remains unclear. Extracellular electrical pulses activate both excitatory and inhibitory neurons ^43–46^, and the net effect likely depends on local excitatory-inhibitory balance, which may be pathology- and localization-specific and difficult to predict a priori.

These findings add to a growing literature suggesting that stimulation-induced changes in spike rate track stimulation’s effect on seizure risk. Short trains of 50 Hz stimulation at charge densities similar to ours increased interictal spike rates at the SOZ, consistent with evidence that these parameters carry a high likelihood of seizure induction^4,47^. Conversely, 2 Hz cortical stimulation at lower charge densities decreased spike rates near the SOZ, consistent with chronic stimulation at these parameters reducing seizure frequency^11–13,48^. The spike rate increase we observed with a seizure induction protocol further supports this hypothesis.

An important caveat is that, although the stimulation protocol was intended to increase seizure risk, it may not have done so uniformly. Only 39.5% of patients had a stimulation-induced seizure, and the spike rate increase was similar between those who did and did not. This could be taken as evidence against the hypothesis that spike rate changes reflect stimulation’s effect on seizure risk. However, it is also consistent with the possibility that the protocol increased seizure risk similarly across patients, but those who had a stimulation-induced seizure had a higher baseline seizure risk to begin with (**Supplemental Fig. S10**).

Spike rates increased rapidly with stimulation onset and returned to baseline within approximately 30 seconds of offset, consistent with a transient, reversible effect on seizure risk. The rapid kinetics of this response raise the possibility that interictal spike rates could serve as a real-time biomarker for optimizing neurostimulation parameters — enabling systematic parameter search within a single clinical session.

### Mesial temporal lobe stimulation causes a disproportionate increase in spike rates, independent of epilepsy localization

Although stimulation-evoked spike rate changes may track seizure risk, their magnitude does not localize the seizure onset zone. The mesial temporal lobe showed the largest stimulation-evoked spike rate increases, independent of epilepsy localization. This suggests that the disproportionate increase in spiking in the mesial temporal lobe reflects intrinsic tissue hyperexcitability rather than a property of the epileptic network — and therefore that a stimulation-evoked increase in spike rates at mesial temporal sites does not imply mesial temporal lobe epilepsy localization^18,51^.

This stands in apparent contrast to multiple studies that find stimulation-induced *seizures* at mesial temporal sites carry moderate specificity for epilepsy localization and laterality^24,52^. These results may nonetheless be compatible if we consider that patients with mesial temporal lobe epilepsy have a higher baseline probability of seizure formation from the epileptic lobe. Stimulation could increase that probability similarly in epileptic and non-epileptic mesial temporal tissue — reflected in the comparable spike rate increases across epilepsy localizations — but cross the seizure threshold only in patients with mesial temporal lobe epilepsy. Consistent with this, as noted above, stimulation-induced seizures in our cohort were not preceded by a greater stimulation-evoked spike rate increase; if stimulation increases excitability similarly regardless of epilepsy localization, no such difference would be expected. A practical implication for surgical planning is that stimulation-evoked spike rate increases at mesial temporal sites should not be weighted as localizing evidence without accounting for the anatomical location of the stimulation target.

### Stimulation-evoked spike rate changes do not improve SOZ localization beyond baseline spike rate

A model incorporating stimulation-induced spikes achieved moderate accuracy in localizing the seizure onset zone (SOZ), consistent with prior studies finding that delayed responses localize the SOZ^15–17^. However, baseline spike rates alone performed comparably, suggesting that stimulation-induced spikes do not add localizing information beyond spikes in the resting state. This is consistent with our finding that stimulation-evoked spike rate increases at mesial temporal sites are independent of epilepsy localization, and together suggest these increases reflect regional tissue excitability rather than a property of the epileptic network.

Whether this null result extends to other stimulation-evoked features — such as high frequency oscillations^53^, cortico-cortical evoked potential (CCEP) amplitudes^54^, and evoked gamma power^55,56^, which differ from acute spike rate changes in their putative mechanisms — remains an open question. Our results argue against performing stimulation solely to elicit spikes for SOZ localization, at least using the features examined here.

### Stimulation alters spike morphology, implicating recruitment of distinct spike generators

Stimulation had a small but significant effect on spike morphology, producing spikes that were broader and higher amplitude. Whether spikes were recorded during or between stimulation explained only 0.36% of total morphological variance; the largest source of variance (21.3%) was recording electrode, consistent with spike morphology reflecting the local neuronal population surrounding each contact. At the local field potential level, spike shape depends on the number of neurons contributing coherent transmembrane currents, their temporal synchrony, and the distance from active sources to the recording contact^38,57,58^. The small but significant increases in amplitude and width are consistent with recruitment of additional neurons into the epileptic discharge — producing a larger and more temporally dispersed response — rather than simply activating the same neurons more frequently. This is consistent with the clinical belief that higher charge density depolarizes a progressively larger neuronal population, so that stimulation-induced seizures and functional responses may originate in regions beyond the direct stimulation site^59^.

### Limitations

Although the scale of our data necessitated automated spike detection, automated detection is inherently imperfect. Sensitivity and specificity were similar across pre-, during-, and post-stimulation periods, mitigating the risk that detection bias could explain the observed increase in spike rates with stimulation. Second, we did not systematically account for state-dependent factors known to modulate interictal spiking, including sleep–wake stage, anti-seizure medication levels, and time since implant. Because each stimulation period served as its own control with paired pre- and post-stimulation epochs, it is unlikely that temporal trends in these factors drove our findings. Third, the designation of mesial temporal lobe epilepsy and the seizure onset zone was a clinical determination, without confirmation by surgical outcomes. Fourth, this is a single-center retrospective cohort study of one set of stimulation parameters; external, multicenter validation is needed to assess generalizability. Our analysis did not account for structural or functional connectivity, which may mediate stimulation effects at distant sites beyond what simple current spread would predict^6,49,50^. Finally, we only studied stimulation with a single parameter set. Explicit comparison of multiple parameter sets is needed to understand which parameters, and under what conditions, tend to increase vs. reduce spike rates.

## Conclusions

Low-frequency cortical stimulation delivered as part of a seizure induction protocol produces a spatially constrained increase in spike rate in nearby high-spiking tissue and at the seizure onset zone, with a stronger effect in the mesial temporal lobe, and subtly reshapes spike morphology in a manner consistent with recruiting a broader neuronal population. The magnitude of this response did not localize the seizure onset zone beyond the baseline spike rate, but it did track the expected effect of stimulation on seizure risk. Our findings argue against using stimulation-evoked spikes for localization, but support the acute change in spike rate as a biomarker of stimulation’s effect on seizure risk to guide parameter selection for treating drug-resistant epilepsy.

## Data availability

All code used to perform analyses, along with an intermediate dataset, is available at https://github.com/penn-cnt/stim-responses. Complete patient iEEG data are available at iEEG.org, which users can access free of charge. All patients are part of the HUP_Intracranial_Data project at iEEG.org; data are available on request.

