## Supplemental Materials for "Regional excitability, not epileptic pathology, drives stimulation-evoked interictal spike increases"

### Supplement

#### Methods

##### Electrode registration

The post-implant head CT was co-registered to the pre-implant T1-weighted MRI and segmented using ANTsPyNet and iEEG-recon[1]. Electrodes were assigned to the nearest region label in the Desikan–Killiany–Tourville (DKT) atlas[2].

##### Spike detection parameter tuning and validation

We performed a hyperparameter search to optimize our previously validated automated spike detector for stimulation recordings. We sampled 1,000 hyperparameter combinations and evaluated each on a labeled training set of 350 50-second clips, sampled across stimulation/recording channel pairs and consisting of 10 s pre-, 30 s during-, and 10 s post-stimulation windows. Clips were sampled across stimulation/recording channel pairs drawn from the patients included in the analysis (n=43). Within each patient, clips were restricted to those in which a first unoptimized pass of the detector identified at least 3 spikes during the 50 s window, ensuring annotated clips contained enough events to discriminate hyperparameter sets. Spikes in these clips were independently labeled by a board-certified epileptologist (E.C.C.) blinded to automated detections. Example spike annotations are included in **Supplemental Fig. S1**. We selected the hyperparameter set that maximized recall, prioritizing detection sensitivity given that subsequent analyses depended on detecting an adequate number of spikes to detect differences across stimulation conditions. The detector applies four hyperparameters that together define what counts as a spike. *tmul* is a relative threshold multiplier: a candidate spike's amplitude must exceed *tmul* × the median absolute amplitude computed in a window surrounding the spike. *absthresh* (μV) is a fixed lower amplitude bound that the spike must exceed regardless of local background, preventing small fluctuations in low-activity segments from passing the relative threshold. *sur_time* (s) is the half-width of the window used to compute the local median amplitude that defines the relative threshold. *too_high_abs* (μV) is an upper amplitude bound used to reject artifact (e.g., movement, residual stimulation artifact). The hyperparameter set selected by the search was tmul = 23.0, absthresh = 150 μV, sur_time = 50 s, and too_high_abs = 2000 μV. To ensure performance generalized beyond the training set, we evaluated the final detector on a held-out test set of 150 50-second clips, computing sensitivity and precision separately for the pre-, during-, and post-stimulation windows of each clip and using a repeated-measures ANOVA to test for bias across the three windows. Visual review of stimulation trials shows minimal false positives (**Supplemental Fig. S2**).

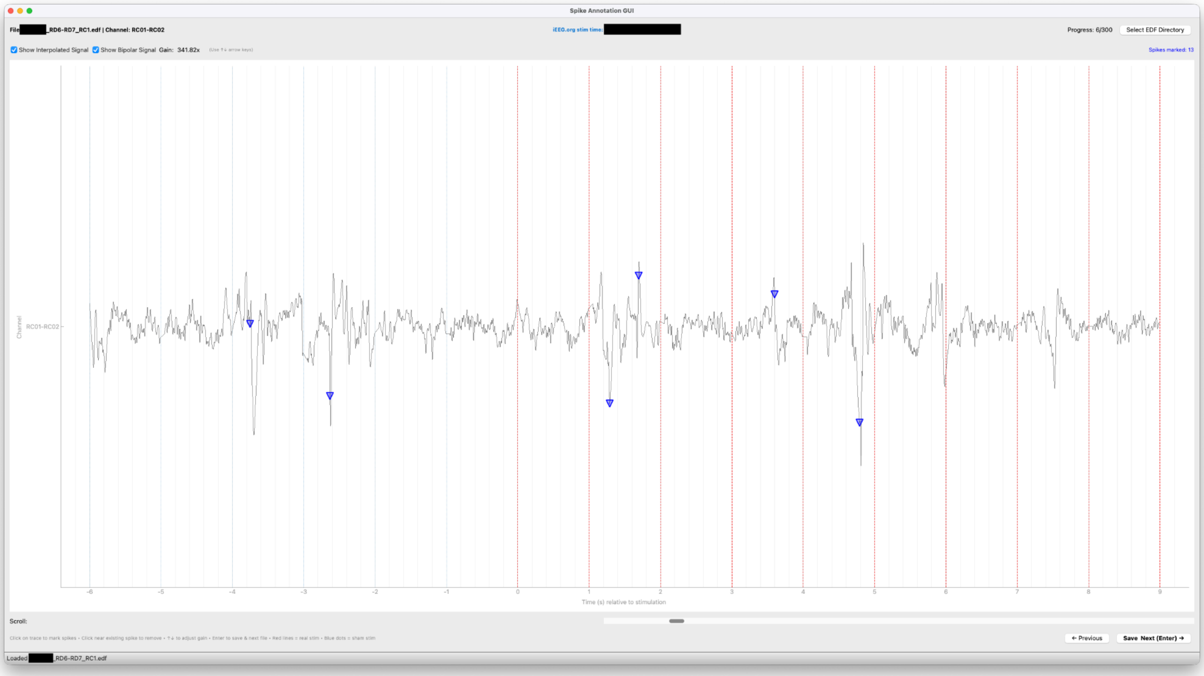


**Supplemental Figure S1. Example clinician annotation of interictal spikes using our GUI.** Blue triangles indicate clinician annotations of spikes.

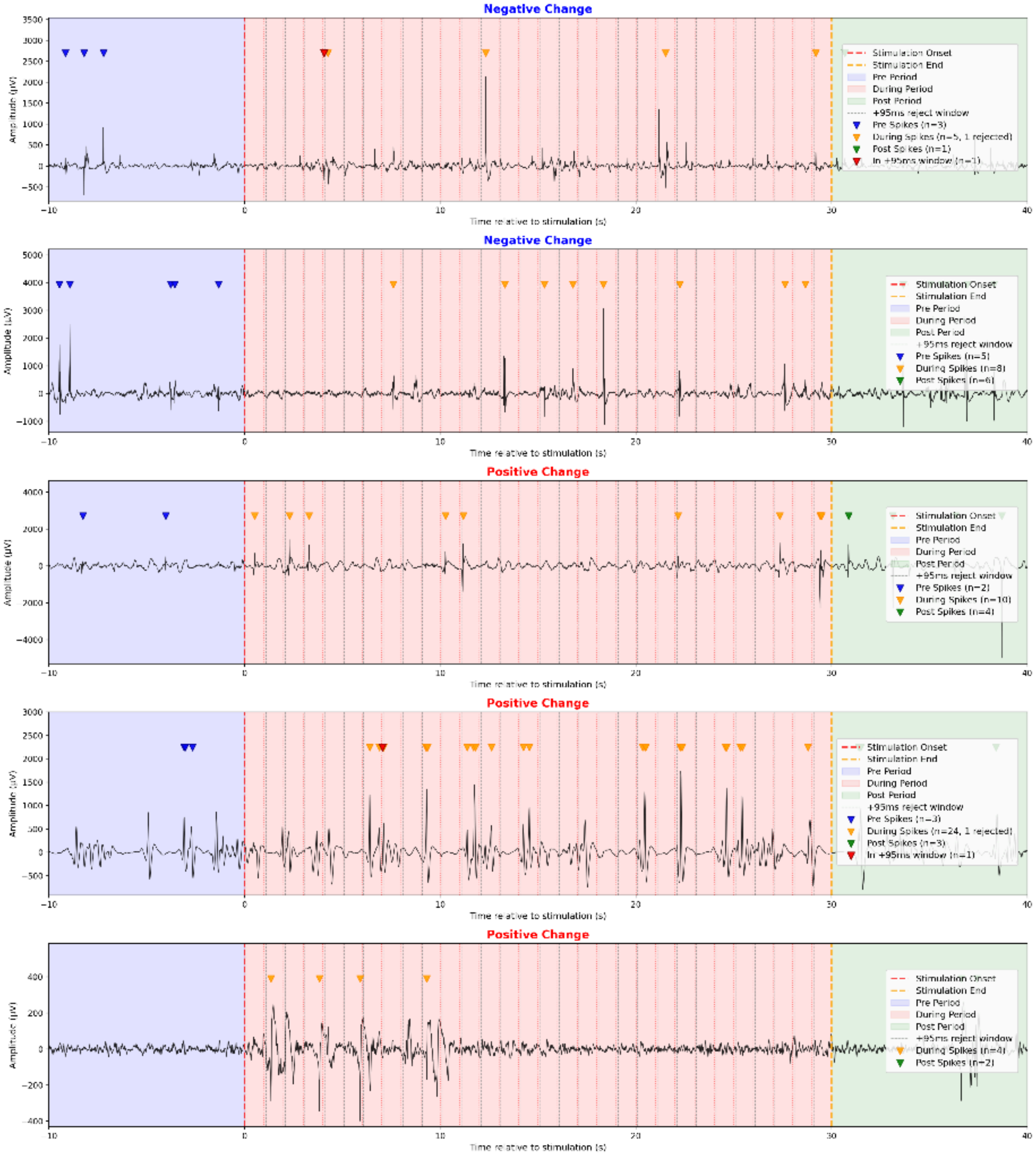
Negative Change
Amplitude (µV)
Time relative to stimulation (s)
Stimulation Onset
Stimulation End
Pre Period
During Period
Post Period
+95ms reject window
Pre Spikes (n=3)
During Spikes (n=5, 1 rejected)
Post Spikes (n=1)
In +95ms window (n=1)
Negative Change
Amplitude (µV)
Time relative to stimulation (s)
Stimulation Onset
Stimulation End
Pre Period
During Period
Post Period
+95ms reject window
Pre Spikes (n=5)
During Spikes (n=8)
Post Spikes (n=6)
Positive Change
Amplitude (µV)
Time relative to stimulation (s)
Stimulation Onset
Stimulation End
Pre Period
During Period
Post Period
+95ms reject window
Pre Spikes (n=2)
During Spikes (n=10)
Post Spikes (n=4)
Positive Change
Amplitude (µV)
Time relative to stimulation (s)
Stimulation Onset
Stimulation End
Pre Period
During Period
Post Period
+95ms reject window
Pre Spikes (n=3)
During Spikes (n=24, 1 rejected)
Post Spikes (n=3)
In +95ms window (n=1)
Positive Change
Amplitude (µV)
Time relative to stimulation (s)
Stimulation Onset
Stimulation End
Pre Period
During Period
Post Period
+95ms reject window
During Spikes (n=4)
Post Spikes (n=2)

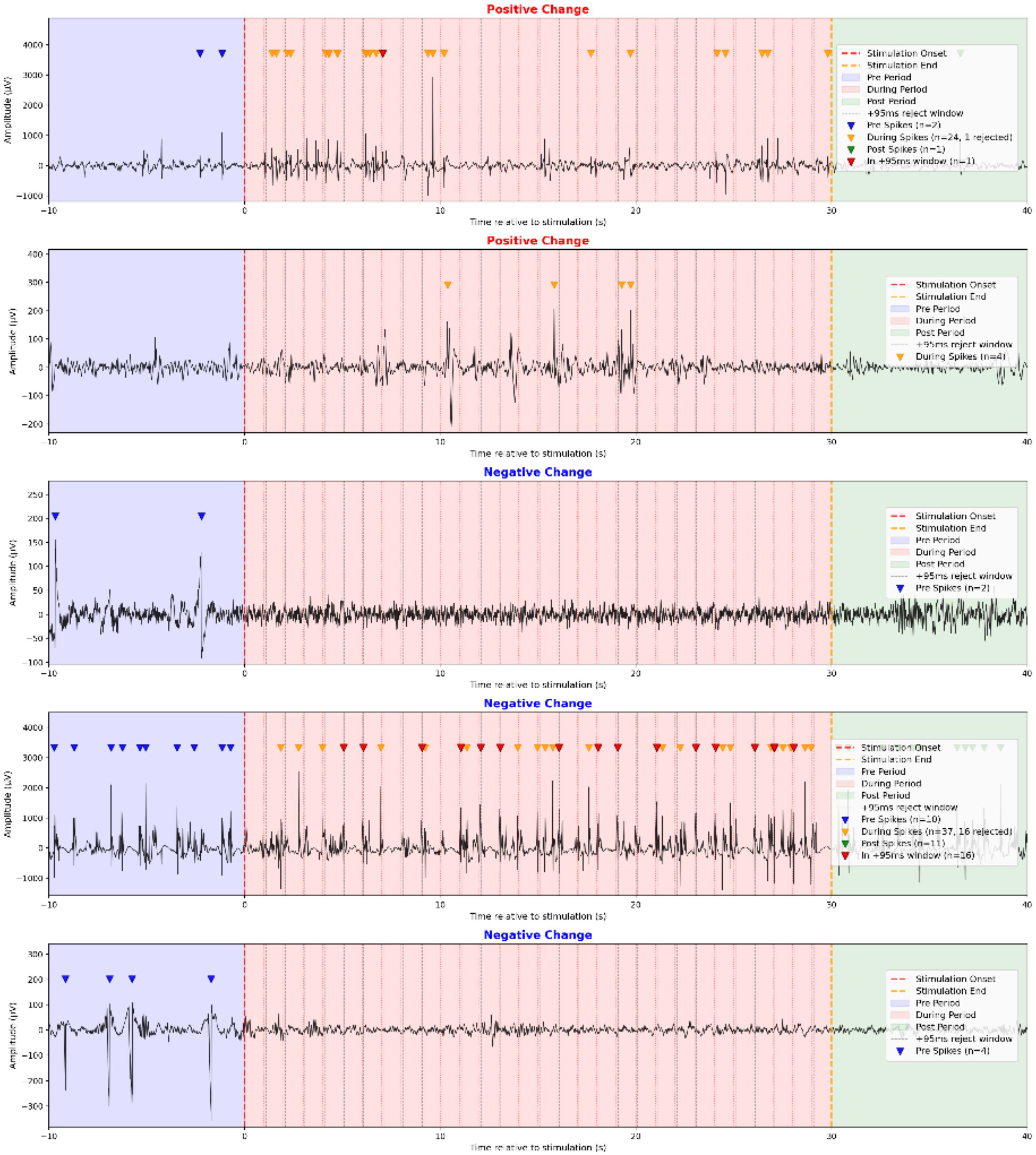


**Supplemental Figure S2. Ten random stimulation trials and their automatic spike detection annotations.** The blue shaded window is the pre-stimulation period, the pink shaded window is the during-stimulation period (with stimulation times marked as vertical lines), and the green shaded window is the post-stimulation period. The title above each subplot denotes whether the detected spike rate was higher during stimulation (positive change) or during the inter-stimulation period (negative change). Blue triangles mark pre-stimulation spike detections, green triangles mark post-stimulation spike detections, orange triangles mark during-stimulation spike detections, and red triangles mark detections that were excluded because they occurred with 95 ms after a stimulation (or sham stimulation, in the inter-stimulation period).

#### Spike Morphology Feature Computation

For each detected spike, we computed eight morphological features describing both the spike body and the after-going slow wave, following our previously published method[3]. After identifying the spike peak (P), we located the local minimum or maximum opposite of P within ±75 ms to define the spike start (L) and spike end (R), based on spike polarity. The slow wave end point (S) was defined as the second zero-crossing in the signal between 200 and 500 ms after R.

From these four feature points, we computed the eight features used as inputs to the PERMANOVA: sharpness, computed as the average of the absolute slope on the rising and decay halves of the spike; width, the duration between L and R; line length, the cumulative sum of absolute amplitude differences between consecutive samples in the spike body; slow wave width, the duration between R and S; slow wave amplitude, the absolute amplitude difference between the slow wave extremum and the baseline; rise slope, $|P - L|$ divided by the duration from L to P; decay slope, $|P - R|$ divided by the duration from P to R; and average amplitude, the mean of the rising amplitude ($|P - L|$) and decay amplitude ($|R - P|$). We removed rising amplitude and decay amplitude, since they were the only features that were highly correlated and could be captured by a single average amplitude feature, consistent with prior research[3].

#### PERMANOVA Stratified Permutation Test

To test the effect of stimulation condition (during vs. inter-stimulation) on multivariate spike morphology while accounting for nuisance sources of variance, we used a stratified permutation MANOVA (PERMANOVA). Pairwise Euclidean distances between spike morphology vectors were converted to a Gower-centered Gram matrix via double-centering, which reformulates pairwise distances as inner products and enables a sums-of-squares decomposition. Variance was partitioned sequentially (Type I SS), entering terms in the order: patient identity, stimulation channel, recording channel, and stimulation condition. This ordering ensures that between-patient differences and electrode-level effects are accounted for before testing the effect of condition.

To assess the significance of the condition effect, we performed a stratified permutation test. Condition labels were shuffled only within each patient, preserving patient-level structure while destroying the within-patient during versus inter-stimulation signal. For each of 999 permutations, SS(condition) was recomputed holding the nuisance hat matrix fixed, and a pseudo-F statistic was calculated. The p-value was defined as the proportion of permuted pseudo-F values greater than or equal to the observed pseudo-F. Effect size was reported as pseudo-$R^2$, defined as SS(condition) divided by SS(total), quantifying the proportion of residual multivariate variance attributable to stimulation condition after removing patient and electrode contributions.

#### Training of the Machine Learning Algorithm for SOZ Classification

We trained a random forest classifier to label channels as belonging to the SOZ within a nested, two-loop cross-validation framework to avoid bias and information leakage. The outer loop consisted of leave-one-out cross-validation (LOOCV) at the patient level: in each iteration, the classifier was trained on all patients except one held-out patient, and performance was evaluated on the held-out patient. The inner loop consisted of cross-validated model selection within the training set, in which we performed a grid search over random forest hyperparameters. Because SOZ channels comprised the minority class (<10% true labels), we addressed class imbalance by applying imbalanced-learn's synthetic minority over-sampling technique (SMOTE)[4,5] only within the inner-loop training folds; resampling was never applied to validation folds or to the held-out patient in the outer loop. The final model used the hyperparameter set selected in the inner loop and was then fit on the full outer-loop training set before generating predictions for the held-out patient. This procedure was repeated until each patient served once as the outer-loop test case. AUC and average precision were computed using each patient's out-of-fold prediction probabilities, and DeLong's method was used to compare AUCs between the full and baseline-only models.

#### Alternative Definition of Mesial Temporal Lobe Epilepsy

We tested whether our findings were sensitive to how we defined mesial temporal lobe epilepsy (MTLE). In the main analysis, a patient was classified as MTLE if any clinically determined seizure onset involved the mesial temporal lobe. To assess whether this definition was too inclusive, we re-ran the analysis using a stricter definition: patients were classified as MTLE only if all clinically determined seizure onsets were localized to the mesial temporal lobe. We then compared the change in spike rate during MTL stimulation between patients meeting this stricter criterion (n = 17) and those who never had a mesial temporal seizure onset (n = 21).

### Results

#### Excluded patients had similar demographic characteristics but lower 24-hour baseline spike rates

15 of 62 patients were excluded due to a low spike detector positive predictive value (PPV; <70%) on the spike-detection validation set. **Supplemental Fig. S3** summarizes patient exclusion criteria. To assess whether these exclusions introduced bias into the cohort, we compared demographic and clinical characteristics between excluded (n = 15) and included (n = 43) patients (**Supplemental Table S1**). Groups did not differ across age at onset, age at implant, sex, and MRI lesionality. Detected spike rates during the 24-hour baseline period were lower in excluded than in included patients (mean ± SD: 0.228 ± 0.23 spikes/min vs. 0.545 ± 0.76 spikes/min; Mann–Whitney p = 0.016; **Supplemental Fig. S4**). This is consistent with lower-spike-density recordings yielding fewer true positives at a fixed false-positive rate, reducing the achievable PPV.

| | Good | Bad |
|---|---|---|
| Total: N (current) | 43 | 15 |
| Female: N (%) | 23 (53.5%) | 7 (46.7%) |
| Age at onset in years: median (range) | 22.0 (1.0 - 61.0) | 20.5 (2.0 - 49.0) |
| Age at implant in years | 39.7 (21.1 - 64.6) | 36.5 (21.1 - 58.0) |
| MRI lesional | 16 (37.2%) | 6 (40.0%) |
| Unilateral SOZ | 32 (74.4%) | 11 (73.3%) |
| Temporal SOZ | 27 (62.8%) | 11(73.3%) |
| # with seizure induced by stim | 17 (39.5%) | 5 (33.3%) |
| Total # of induced seizures | 18 | 6 |
| Resection or ablation | 22 (51.2%) | 9 (60.0%) |
| Baseline spike rate: mean ± SD | 0.55 ± 0.76 | 0.23 ± 0.24 |

**Supplemental Table S1. Demographic and clinical characteristics for excluded and included patients.**

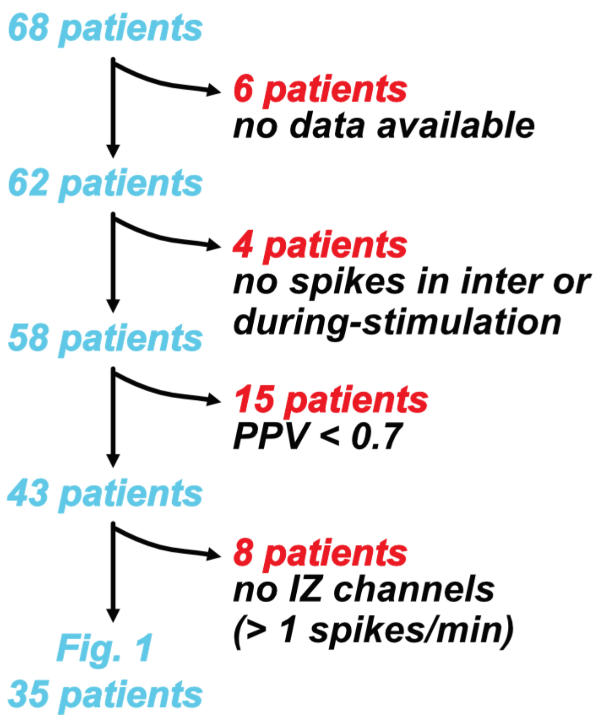


**Supplemental Figure S3. Flowchart describing patient dropoff during main analysis.**

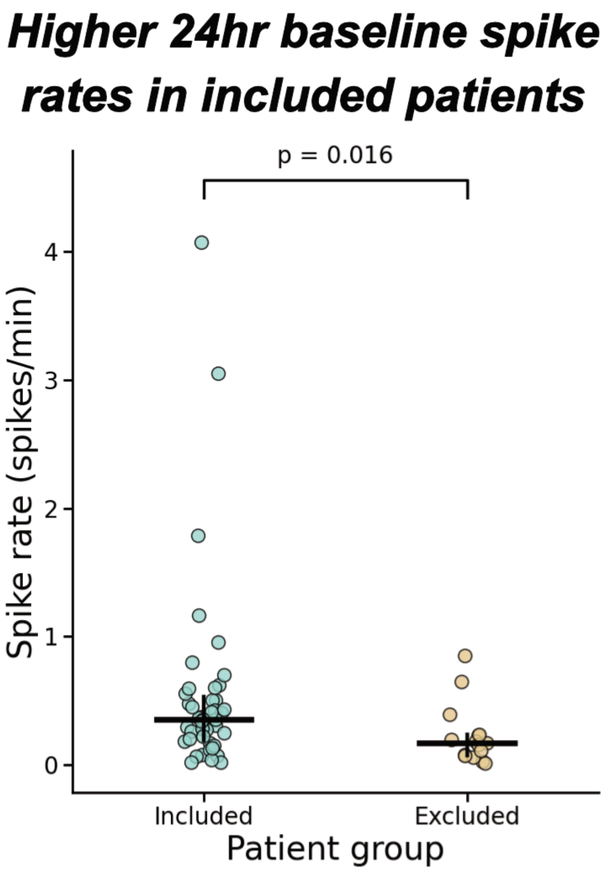


**Supplemental Figure S4. 24-hour baseline spike rates for excluded and included patients.** Each dot represents one patient; black bars indicate the cohort mean. Excluded patients had significantly lower mean spike rates (Mann–Whitney p = 0.016).

#### Spike detector sensitivity and precision are stable

We assessed whether stimulation introduced systematic bias into automated spike detection. On the held-out test set (n = 150 clips), sensitivity (mean ± SEM: pre = 0.67 ± 0.04, during = 0.64 ± 0.04, post = 0.71 ± 0.04) and precision (pre = 0.68 ± 0.04, during = 0.72 ± 0.04, post = 0.74 ± 0.04) did not differ across stimulation windows (repeated-measures ANOVA; sensitivity: F = 0.994, p = 0.371; precision: F = 1.010, p = 0.365; **Supplemental Fig. S5**). These results support that observed changes in spike rate between during- and inter-stimulation windows reflect true changes in neural activity rather than a detection bias introduced by stimulation artifact.

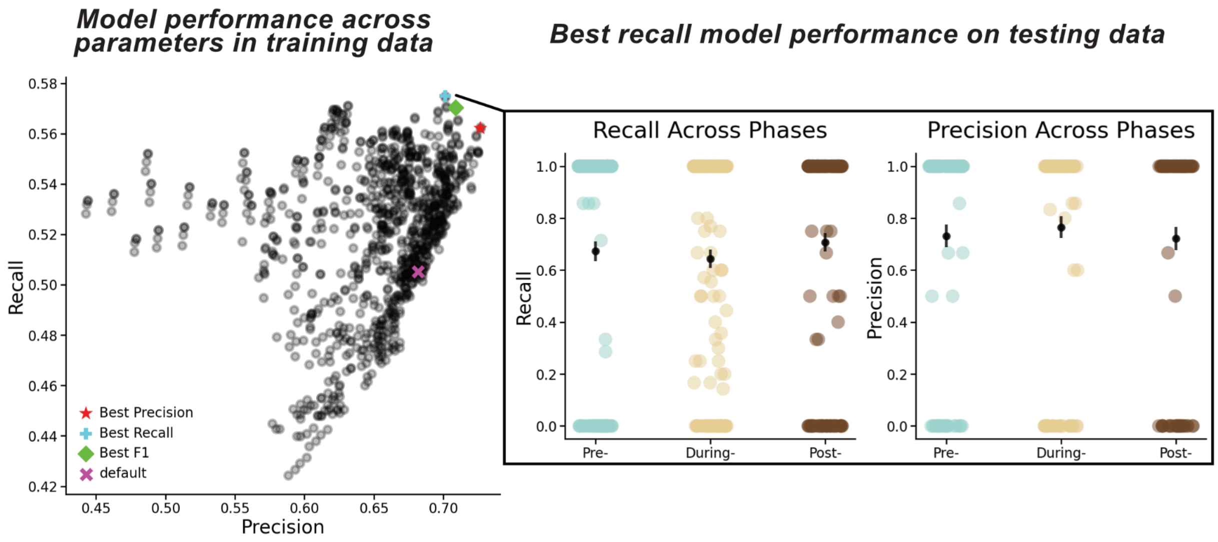


**Supplemental Figure S5. Spike detector performance across stimulation windows.** Recall/sensitivity (left) and precision/positive predictive value (right) of the final spike detector for the pre-, during-, and post-stimulation windows of the held-out test set (n = 150 clips, gold standard was clinician annotations of interictal spikes). Bars indicate the mean ± SEM across clips. Neither metric differed significantly across windows (repeated-measures ANOVA; sensitivity F = 0.994, p = 0.371; precision F = 1.010, p = 0.365). Each dot is a single 50-s stimulation period. Recall and precision often clustered at 0 and 1 because many of the 50-s periods had a low number of spikes in each window, leading to 100% sensitivity if they were all correctly detected, and 0% sensitivity if none were correctly detected, for instance.

#### Inter-stimulation spike rate correlates with the 24-hour baseline spike rate

We assessed whether the inter-stimulation period provided a representative estimate of each patient's resting-state interictal spike activity. For each patient, we computed the channel-wise Spearman correlation between the 24-hour baseline spike rate and the inter-stimulation spike rate. The median (IQR) correlation across patients was 0.57 (0.38 – 0.73; **Supplemental Fig. S6**), indicating that channel-level spike rates were broadly consistent between extended baseline recordings and the brief inter-stimulation epochs used as a within-session control.

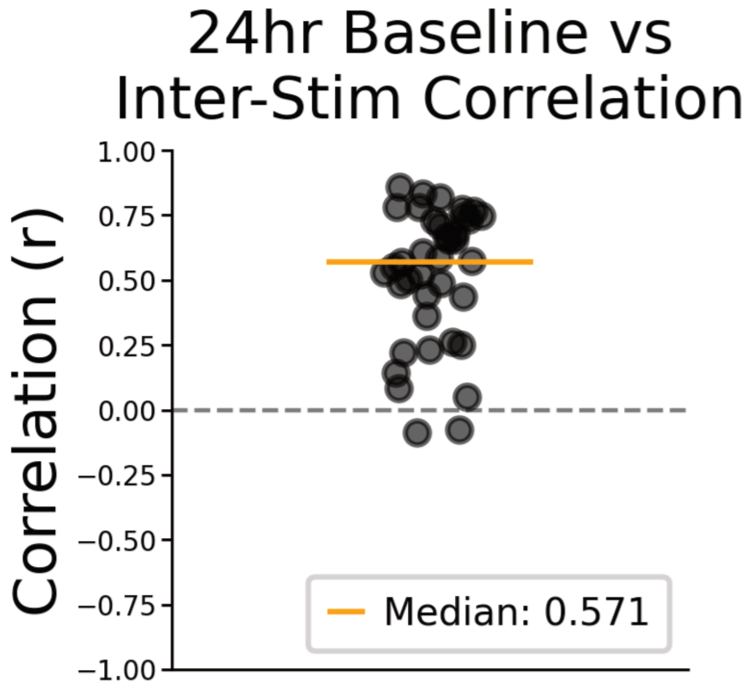


**Supplemental Figure S6. Channel-wise correlation between 24-hour baseline and inter-stimulation spike rate.** Each dot represents one patient's Spearman correlation between channel-level spike rates measured during the 24-hour baseline period and during inter-stimulation epochs. The black bar indicates the median; whiskers indicate the IQR. Median ρ = 0.57 (IQR 0.38 – 0.73).

#### Stimulation-induced spike rate increase replicates at the seizure onset zone

We tested whether the increase in spike rate observed in the irritative zone with nearby stimulation also occurred when restricting the analysis to clinically defined SOZ channels. Across SOZ channels with stimulation delivered within 40 mm (n = 38), the during-stimulation spike rate (median 2.64; IQR 1.14 – 5.86 spikes/min) was significantly higher than the inter-stimulation spike rate (1.91; 0.89 – 4.22 spikes/min; Wilcoxon signed-rank, p = 0.021; Cliff's δ = 0.11; **Supplemental Fig. S7A**). Stratifying by stimulation phase, spike rate increased significantly from pre-stimulation (median 1.16; IQR 0.44 – 4.58 spikes/min) to during-stimulation (2.64; 1.14 – 5.86 spikes/min; Bonferroni-corrected p = 0.023; Cliff's δ = 0.18). The post-stimulation spike rate (1.63; 0.95 – 4.43 spikes/min) was numerically lower than the during-stimulation peak but did not differ significantly from it (Cliff's δ = −0.11; **Supplemental Fig. 7B**). The smaller effect size at the SOZ relative to the irritative zone is consistent with the smaller number of channels meeting strict SOZ criteria, reducing power relative to studying the larger number of channels meeting criteria for inclusion in the irritative zone.

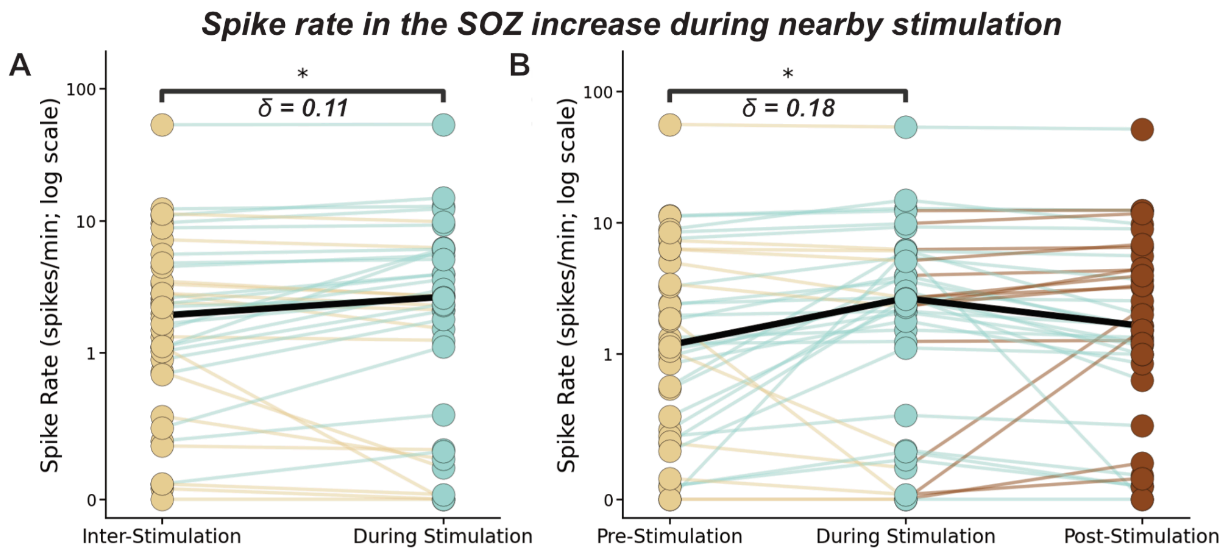


**Supplemental Figure S7. Stimulation-induced increase in spike rate at the SOZ.** (A) SOZ spike rate during inter- and during-stimulation periods (n = 38 patients with both measures). (B) SOZ spike rate stratified by pre-, during-, and post-stimulation windows (same patients). Each dot represents one patient; black lines indicate the cohort median. Effect sizes (Cliff's δ) and Bonferroni-corrected significance levels are shown for each comparison.

#### Spike rate is no longer elevated above baseline by 60–120 s after stimulation offset

We performed a limited, exploratory paired analysis to assess whether residual excitability persisted beyond the immediate post-stimulation window. For each patient, we compared two post-stimulation windows from that patient's *last* stimulation train (0–60 s and 60–120 s post-stimulation) against the trial-averaged pre- and during-stimulation spike rates across all qualifying nearby IZ trials. The analysis was restricted to n = 14 patients with both qualifying nearby IZ trials and a complete 120 s post-stimulation window after their last train (one patient was excluded for lacking a 60–120 s window; **Supplemental Fig. S8**). At this sample size, paired Wilcoxon tests were underpowered for anything but large effects, so we treat these results as exploratory. Trial-averaged spike rate increased from pre-stimulation (median 2.18; IQR 1.52–5.23 spikes/min) to during-stimulation (5.50; 3.15–8.57 spikes/min; Bonferroni-corrected p = 0.024, δ = 0.50). By 60–120 s after the final stimulation train, spike rate (0.23; 0.00–1.26 spikes/min) was significantly lower than the during-stimulation peak (p = 0.010, δ = 0.70). Both post-stimulation windows were numerically below the pre-stimulation baseline.

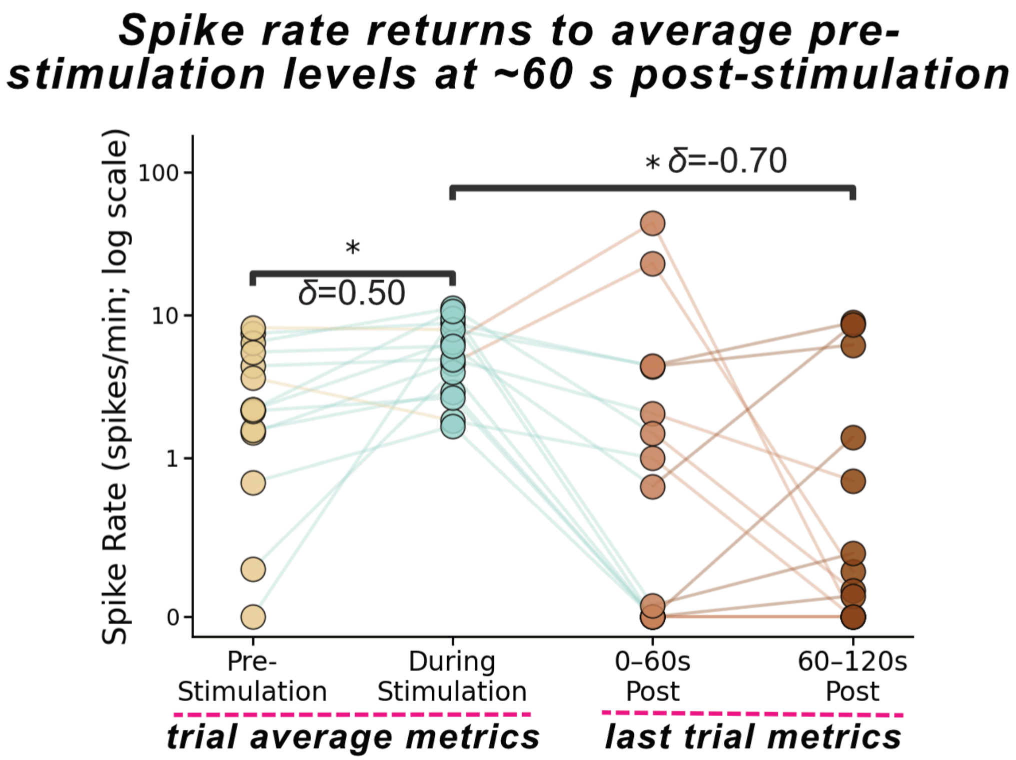


**Supplemental Figure S8. Spike rate returns to pre-stimulation levels by 60–120 s post-stimulation.** Per-patient spike rate (spikes/min; log scale) across pre-stimulation, during-stimulation, 0–60 s post-stimulation, and 60–120 s post-stimulation windows, restricted to patients with qualifying nearby IZ stimulation trials and a complete 60–120 s post-stimulation epoch (n = 14). Each dot represents one patient; lines connect within-patient measurements across windows. Spike rate increased significantly from pre- to during-stimulation (Bonferroni-corrected $p = 0.024$; Cliff's $\delta = 0.50$) and decreased significantly from during-stimulation to 60–120 s post-stimulation ($p = 0.010$; $\delta = -0.70$). *$p < 0.05$.

#### Restricting MTLE classification to patients with only mesial temporal seizures does not change findings

To assess whether our main result — that the change in spike rate during MTL stimulation did not differ between MTLE and non-MTLE patients — was sensitive to how MTLE was defined, we re-ran the analysis using a stricter MTLE definition restricted to patients whose only clinically determined seizure onsets were in the mesial temporal lobe (n = 17), excluding patients with mixed MTL and non-MTL seizure onsets. Patients without any mesial temporal seizure onset (n = 21) served as the comparison group. The change in spike rate during MTL stimulation did not differ significantly between groups (only-MTL: median Δ = 0.26, IQR −0.13 – 0.58 spikes/min; no-MTL: 0.39, −0.02

– 1.68 spikes/min; Mann–Whitney U = 118, p = 0.078; **Supplemental Fig. S9-*ii***). Within-group analyses showed that spike rate increased significantly during MTL stimulation in the no-MTL group (Wilcoxon W = 39, Bonferroni-corrected p = 0.013) but not in the only-MTL group (W = 52, Bonferroni-corrected p = 0.527; **Supplemental Fig. S9-*i***). These results are consistent with the main analysis: the acute excitatory response to MTL stimulation is not stronger in patients with mesial temporal lobe epilepsy.

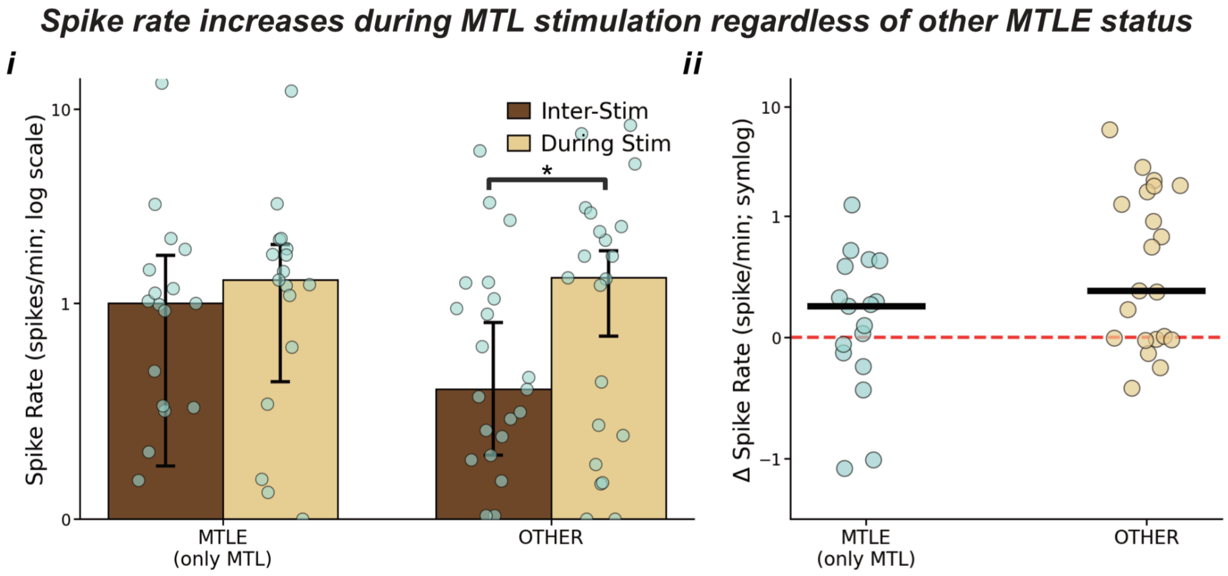


**Supplemental Figure S9. Change in spike rate during MTL stimulation under a stricter MTLE definition.** (*i*) Spike rate during inter- and during-stimulation for patients with only mesial temporal seizures (only-MTL; n = 17) and patients without any mesial temporal seizure onset (no-MTL; n = 21). (*ii*) Distribution of Δ spike rate per patient. Black bars indicate the median; whiskers indicate the IQR. Within-group Wilcoxon tests showed a significant increase in the no-MTL group (Bonferroni-corrected p = 0.013) but not the only-MTL group (Bonferroni-corrected p = 0.527). The between-group comparison was not significant (Mann–Whitney p = 0.078).

#### Hypothesized mechanisms for stimulation-induced seizures

Two mechanisms could explain why LFS induces seizures in some patients but not others. First, stimulation may increase seizure risk equivalently across patients, but only those with high baseline seizure risk cross the seizure threshold (**Supplemental Fig. S10, left**). Second, stimulation may produce a larger increase in excitability in patients who develop seizures, crossing the threshold due to a greater stimulation-induced change rather than a higher starting point (**Supplemental Fig. S10, right**). The absence of a greater stimulation-evoked spike rate increase in patients who had stimulation-induced seizures (**Fig. 3E**) is inconsistent with the second mechanism, and instead supports that seizure induction reflects high baseline seizure risk rather than a disproportionate acute excitatory response.

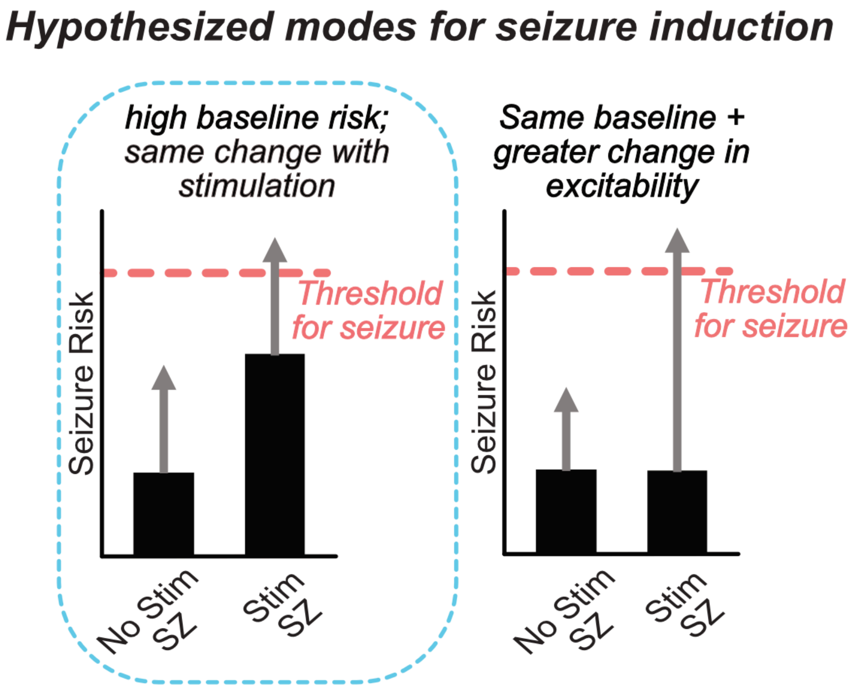


**Supplemental Figure S10. Hypothesized modes for stimulation-induced seizure.** Two mechanisms could explain seizure induction in a subset of patients: equivalent excitability increase across patients with seizures occurring only in those with high baseline risk (left, highlighted), or a disproportionately large excitability increase selectively in patients who develop seizures (right). SZ = seizure.

#### Average precision of SOZ classification is consistent with AUC analysis

We additionally evaluated precision–recall performance of the SOZ localization models. The full classifier achieved an average precision (AP) of 0.204 [95% CI: 0.129–0.326], compared to 0.171 [0.108–0.273] for the baseline-only model (**Supplemental Figure. S11**). Both models substantially exceeded the no-skill prevalence of 0.07, with peak precision near 0.30–0.35 achieved at low-to-moderate recall before declining at higher recall values. Confidence intervals overlapped broadly across the full recall range, consistent with the AUC analysis showing no meaningful improvement in SOZ localization when stimulation-evoked spike modulation features were added to the 24-hour baseline spike rate.

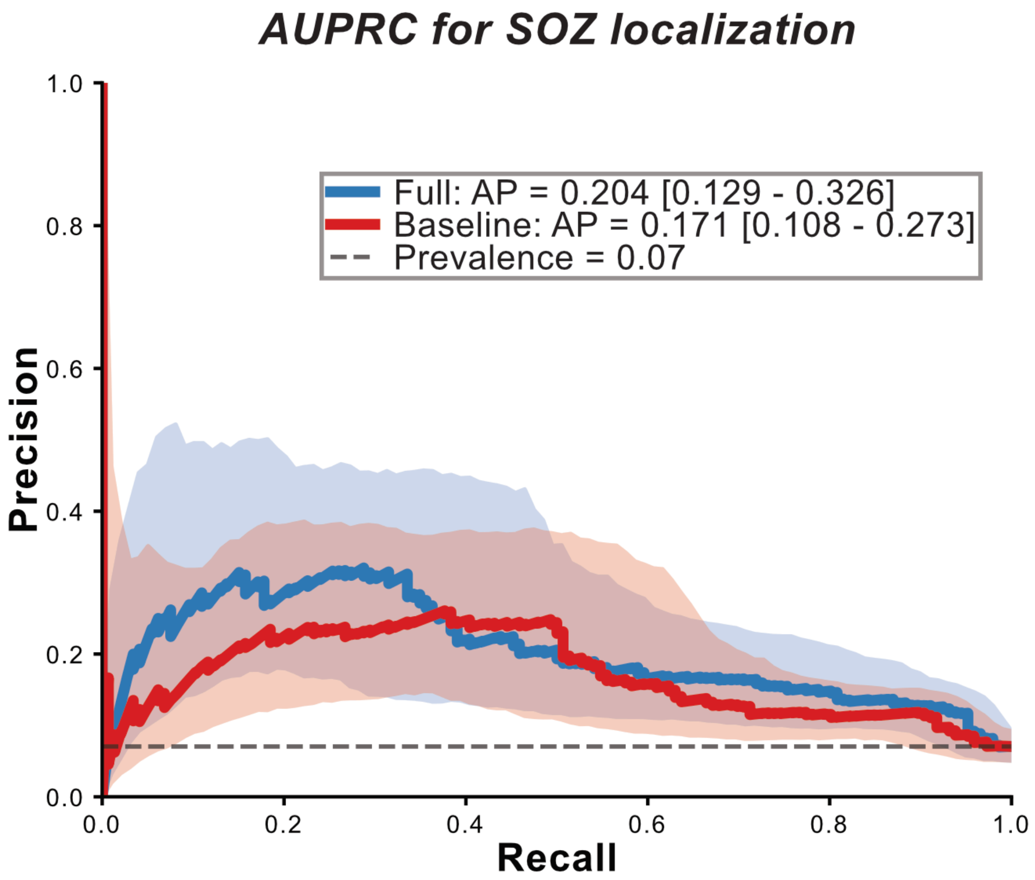


**Supplemental Figure S11. Precision–recall curves for SOZ localization.** Precision–recall curves are shown for the full classifier (blue; 24-hour baseline spike rate, outward modulation, and inward modulation) and the baseline-only model (red; 24-hour spike rate alone). Solid lines represent mean performance across cross-validation folds; shaded regions indicate 95% confidence intervals. The dashed gray line denotes the no-skill classifier (prevalence = 0.07). Average precision (AP) with 95% confidence intervals is reported in the legend for each model.

#### All spike rate measures localize the SOZ with equivalent performance

To determine whether the timing of spike rate measurement relative to stimulation affects SOZ localization, we compared the discriminative performance of three per-channel spike rate measures: the inter-stimulation rate, the during-stimulation rate, and the 24-hour baseline rate. All three measures exceeded chance, with AUCs of 0.701 [95% CI: 0.627–0.763], 0.766 [0.703–0.824], and 0.778 [0.720–0.832], respectively (**Supplemental Fig. S12A**). No pairwise difference in AUC was statistically significant (inter-stimulation vs. during-stimulation: p = 0.75; inter-stimulation vs. 24-hour baseline: p = 0.76; during-stimulation vs. 24-hour baseline: p = 0.95; DeLong test).

Precision–recall analysis was consistent with this finding: average precision was 0.146 [0.094–0.220], 0.195 [0.124–0.307], and 0.179 [0.115–0.284] for the inter-stimulation, during-stimulation, and 24-hour baseline rates, respectively, with confidence intervals overlapping broadly across all comparisons (**Supplemental Fig. S12B**; no-skill prevalence = 0.06). These results indicate that SOZ localization performance is not sensitive to which recording epoch is used to estimate per-channel spike rate, and that neither stimulation-period nor post-stimulation spike dynamics carry additional localizing information beyond the 24-hour baseline rate.

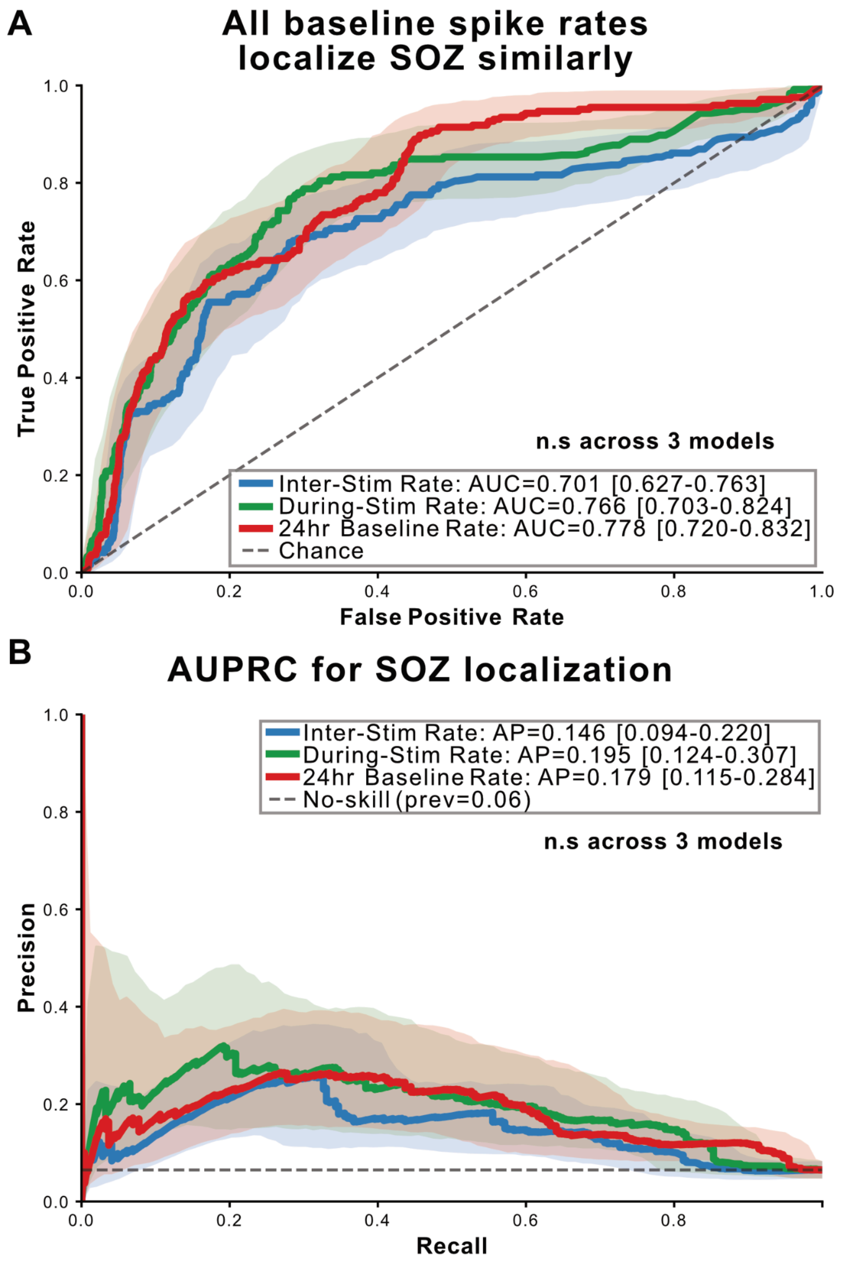

**Supplemental Figure S12. All spike rate measures localize the SOZ with equivalent performance.** (A) Receiver operating characteristic (ROC) curves for SOZ localization using per-channel inter-stimulation spike rate (blue), during-stimulation spike rate (green), and 24-hour baseline spike rate (red). AUC with 95% confidence intervals is shown for each model; the dashed gray line indicates chance performance. Pairwise DeLong test p-values are reported below the panel. (B) Precision–recall curves for the same three models. Average precision (AP) with 95% confidence intervals is shown in the legend; the dashed gray line indicates the no-skill classifier (prevalence = 0.06). Solid lines represent mean performance; shaded regions indicate 95% confidence intervals across cross-validation folds.
